# Human Applications of Transcranial Temporal Interference Stimulation: A Systematic Review

**DOI:** 10.1101/2025.05.16.25327804

**Authors:** Ilya Demchenko, Ishaan Tailor, Sina Chegini, Haochen Yu, Fatemeh Gholamali Nezhad, Alice Rueda, Anne Kever, Sridhar Krishnan, Abhishek Datta, Jed A. Meltzer, Simon J. Graham, Tom A. Schweizer, Sumientra Rampersad, Edward S. Boyden, Ines R. Violante, Robert Chen, Andres M. Lozano, Venkat Bhat

**Affiliations:** Interventional Psychiatry Program, St. Michael’s Hospital – Unity Health Toronto, 193 Yonge Street, Toronto, ON M5B 1M4, Canada; Institute of Medical Science, Temerty Faculty of Medicine, University of Toronto, 6 Queen’s Park Crescent, Toronto, ON M5S 3H2, Canada; Institute for Biomedical Engineering, Science and Technology (iBEST), Keenan Research Centre for Biomedical Science, St. Michael’s Hospital – Unity Health Toronto, 209 Victoria Street, Toronto, ON M5B 1X3, Canada; BARLO Multiple Sclerosis Centre and Division of Neurology, St. Michael’s Hospital – Unity Health Toronto, 30 Bond Street, Toronto, ON M5B 1W8, Canada; Department of Psychology, Faculty of Arts & Science, University of Toronto, 100 St. George Street, Toronto, ON M5S 3G3, Canada; Department of Electrical, Computer, and Biomedical Engineering, Toronto Metropolitan University, Toronto, 350 Victoria Street, ON M5B 2K3, Canada; Research and Development, Soterix Medical, Inc., 1480 US-9, Woodbridge, NJ 07095, United States; Department of Biomedical Engineering, City College of New York, 160 Convent Avenue, New York, NY 10031, United States; Rotman Research Institute, Baycrest Hospital, 3560 Bathurst Street, Toronto, ON M6A 1W1, Canada; Physical Sciences Platform, Sunnybrook Research Institute, 2075 Bayview Avenue, Toronto, ON M4N 3M5, Canada; Department of Medical Biophysics, Temerty Faculty of Medicine, University of Toronto, 101 College Street, Toronto, ON M5G 2C4, Canada; Keenan Research Centre for Biomedical Science, St. Michael’s Hospital – Unity Health Toronto, 209 Victoria Street, Toronto, ON M5B 1X3, Canada; Neuroscience Research Program, St. Michael’s Hospital – Unity Health Toronto, 209 Victoria Street, Toronto, ON M5B 1X3, Canada; Division of Neurosurgery, Department of Surgery, Temerty Faculty of Medicine, University of Toronto, 149 College Street, Toronto, ON M5T 1P5, Canada; Department of Physics, University of Massachusetts Boston, 100 William T Morrissey Boulevard, Boston, MA 02125, United States; Department of Electrical and Computer Engineering, Northeastern University, 100 Forsyth Street, Boston, MA 02115, United States; Department of Brain and Cognitive Sciences, Media Arts and Sciences, and Biological Engineering, McGovern Institute for Brain Research and Koch Institute for Integrative Cancer Research, Massachusetts Institute of Technology, 77 Massachusetts Avenue, Cambridge, MA 02139, United States; Howard Hughes Medical Institute, 4000 Jones Bridge Road, Chevy Chase, MD 20815, United States; School of Biomedical Engineering & Imaging Sciences, King’s College London, Strand, London WC2R 2LS, United Kingdom; School of Psychology, Faculty of Health and Medical Sciences, University of Surrey, 30 Priestley Road, Guildford GU2 7YH, United Kingdom; Krembil Brain Institute, Toronto Western Hospital – University Health Network, 135 Nassau Street, Toronto, ON M5T 1M8, Canada; Edmond J. Safra Program in Parkinson’s Disease, Morton and Gloria Shulman Movement Disorders Clinic, Toronto Western Hospital – University Health Network, 399 Bathurst Street, Toronto, ON M5T 2S6, Canada; Division of Neurology, Department of Medicine, Temerty Faculty of Medicine, University of Toronto, 6 Queen’s Park Crescent West, Toronto, ON M5S 3H2, Canada; Department of Psychiatry, Temerty Faculty of Medicine, University of Toronto, 250 College Street, Toronto, ON M5T 1R8, Canada

**Keywords:** temporal interference, electric stimulation, deep brain stimulation, clinical study, humans, brain, systematic review

## Abstract

**Background:** Many neurological and psychiatric disorders involve dysregulation of subcortical structures. Transcranial temporal interference stimulation (tTIS) is a novel, non-invasive method developed to selectively modulate deep brain regions and associated neural circuits.

**Methods:** A systematic review was conducted to evaluate human applications of tTIS (PROSPERO ID: CRD42024559678). MEDLINE, Embase, APA PsycINFO, CENTRAL, ClinicalTrials.gov, and WHO ICTRP were searched up to December 12, 2024. Studies involving human applications of tTIS were eligible. Methodological quality was appraised using the NIH and modified Oxford Centre for Evidence-Based Medicine tools.

**Results:** Forty-eight records were reviewed (20 published studies, 28 ongoing trials). Of published studies, 16 single-session and 4 multi-session studies assessed safety, mechanistic outcomes, or therapeutic effects of tTIS in 820 participants. Stimulation was most commonly delivered at beta (20 Hz) or gamma (30–130 Hz) envelope frequencies. Neuroimaging studies support target engagement of the motor cortex, basal ganglia, and hippocampus in humans, particularly when stimulation is paired with behavioural tasks. Preliminary clinical findings in small samples demonstrated acute symptom improvements in bradykinesia and tremor within 60 minutes following a single tTIS session in Parkinson’s disease and essential tremor. Reported adverse events across studies were mild (e.g., tingling, itching). Emerging trials increasingly utilize multi-session protocols (2–40 sessions) and are extending tTIS to patients with neurological and psychiatric disorders, particularly epilepsy and depression.

**Conclusions:** Phase 1 studies demonstrate that tTIS is safe, well-tolerated, and capable of engaging deep brain targets in humans. Well-controlled Phase 2 trials are needed to assess its therapeutic potential in patient populations.

**Highlights:**

- tTIS engages the motor cortex, basal ganglia, and hippocampus across human studies
- 20 studies show tTIS is safe and well-tolerated in healthy and clinical cohorts
- One tTIS session improves bradykinesia and tremor in Parkinsonism within 1 hour
- Multi-session trials now test tTIS in epilepsy, depression, and other disorders
- Robust Phase 2 trials are needed to study the efficacy of tTIS in patient populations

## 1. Introduction

Neurological and psychiatric disorders affect nearly a quarter of the global population over the course of a lifetime and account for more than 15% of global Disability-Adjusted Life Years (DALYs) [1,2]. Many of these conditions involve dysregulation of subcortical structures, driving efforts to develop interventions capable of targeting deep brain regions [3]. Invasive deep brain stimulation (DBS) has demonstrated relatively high response rates in treatment-resistant populations (40-70%) but carries inherent risks associated with surgical implantation, including hemorrhage, infection, and hardware-related complications, as well as high costs [3,4]. Despite these drawbacks, DBS remains the most effective neuromodulation approach for deep targets, and non-invasive techniques have yet to match its therapeutic efficacy.

Non-invasive neuromodulation approaches such as transcranial magnetic stimulation (TMS), which is FDA-cleared for multiple indications, and transcranial electrical stimulation (tES) offer safer alternatives with fewer adverse effects. However, these approaches typically yield more modest response rates (33-45%) compared to invasive modalities [5,6]. Their limited spatial selectivity and poor capacity to engage subcortical structures have been identified as possible factors influencing efficacy [7–9], although outcomes are likely multifactorial and also shaped by stimulation parameters, patient characteristics, and protocol design. Transcranial ultrasound is another promising modality with excellent spatial precision for subcortical targets; however, broader clinical adoption has been hindered by technical challenges, including skull-induced acoustic attenuation, complex parameter optimization, and safety concerns such as unintended tissue heating [10–12].

These limitations have prompted growing interest in transcranial temporal interference stimulation (tTIS), a novel non-invasive technique designed to modulate deep brain structures using electric fields [13–15]. tTIS delivers two slightly different high-frequency currents (e.g., 2.00 and 2.01 kHz) that interact to produce an amplitude-modulated field with a kHz carrier frequency and lower-frequency envelope. The field strength distribution at the tTIS carrier frequency is similar to those of its sister technologies transcranial direct (tDCS) and alternating (tACS) current stimulation, broadly affecting superficial cortical regions. However, for tTIS this field is at a much higher frequency, which neurons respond to differently, and potentially not at all. Importantly, the field at the envelope frequency (e.g., 10 Hz) created by interference has a more focal area of peak strength that may occur deep in the brain, depending on the electrode placement [13,16]. This interference pattern enables spatially selective targeting of deep brain regions with minimal co-stimulation of the overlying cortex, a property not achievable with conventional tES [13,17]. Compared with TMS, which induces suprathreshold activity but loses focality with depth, and transcranial ultrasound stimulation, which can reach deep regions but faces uncertainties regarding skull attenuation and utilizes mechanical rather than electrical energy, tTIS offers a unique balance of depth penetration and tolerability [10–12,18,19].

Mechanistically, initial models suggested that neurons respond directly to the amplitude-modulated envelope of the interfering currents, but more recent evidence indicates contributions from network-level interactions and ion channel dynamics [16,20–22]. Importantly, all human tTIS studies to date have used subthreshold stimulation, with electric field strengths far weaker than those applied in DBS and TMS, implying fundamentally distinct mechanisms of action. Furthermore, simulations suggest that achieving suprathreshold tTIS in humans may not be feasible with current technology, as this would require intensities exceeding 38 mA [17], which no existing stimulator can deliver and which would likely be intolerable even at 10 kHz. Suprathreshold effects might only be achievable at much higher carrier frequencies, which are not yet available, or potentially through intracranial electrodes. Although not a replacement for DBS, tTIS may offer a more accessible and lower-risk alternative for patients who are ineligible for invasive interventions. Moreover, it holds promise as a complementary or adjuvant tool alongside pharmacotherapy and DBS, or even as a predictive probe for deep target engagement. This systematic review aims to synthesize current evidence on human applications of tTIS and outline directions for future research.

## 2. Methods

### 2.1. Search Strategy

This systematic review followed the Preferred Reporting Items for Systematic Reviews and Meta-Analyses (PRISMA) guidelines (PROSPERO ID: CRD42024559678) [23]. A comprehensive search of OVID (MEDLINE, Embase, APA PsycINFO) and CENTRAL was conducted on December 12, 2024, with a supplementary PubMed search to capture unindexed publications. Clinical trial records were searched on ClinicalTrials.gov and the World Health Organization International Clinical Trials Registry Platform (WHO ICTRP). The search terms were synonyms of temporal interference and electrical stimulation (**eMethods** in the **Supplement**).

### 2.2. Study Selection

Two reviewers independently conducted first-level (i.e., titles and abstracts) and second-level (i.e., full-text) screening, and discrepancies were resolved through consensus with a third party. Primary research articles and clinical trial records were included in the review if they administered tTIS to human participants (list of eligibility criteria in **eMethods** in the **Supplement**).

### 2.3. Data Extraction and Appraisal of Methodological Quality

Extracted data included bibliographic information, participant characteristics, study design, collected outcomes, stimulation parameters, and results (list of variables in **eMethods** in the **Supplement**). Two reviewers independently assessed the quality of evidence using National Institutes of Health tools for controlled intervention studies and uncontrolled pretest-postest designs, and a rating scheme adapted from the Oxford Centre for Evidence-based Medicine (**eTables S1**-**3** in the **Supplement**) [24].

### 2.4. Statistical Analysis

Analyses were performed in R (v.4.4.2) [25]. Frequency counts were used to summarize study characteristics, stimulation parameters, outcomes, and adverse events (AE). Efficacy was assessed using an adapted classification framework [26]. For studies with individual-level data, Hedge’s *g* and mean differences with 95% confidence intervals (CI) were calculated. AE rates were analyzed using _χ_*²* tests with Yates’s continuity correction (*P* < .05). Due to outcome heterogeneity, meta-analysis was not performed.

## 3. Results

### 3.1. Study Selection

The initial search across all databases yielded a total of 3,769 records (**Figure 1**). After removing 1,575 duplicates, 2,194 records were screened by title and abstract. Eighty underwent full-text review, resulting in 48 included records: 20 published studies/protocols and 28 ongoing clinical trials available between September 2018 and December 2024 (**Figure 2A**). Across the published studies, a total of 820 human participants were enrolled, with an additional 2,303 participants projected to be enrolled in the ongoing clinical trials.

**Figure 1.**
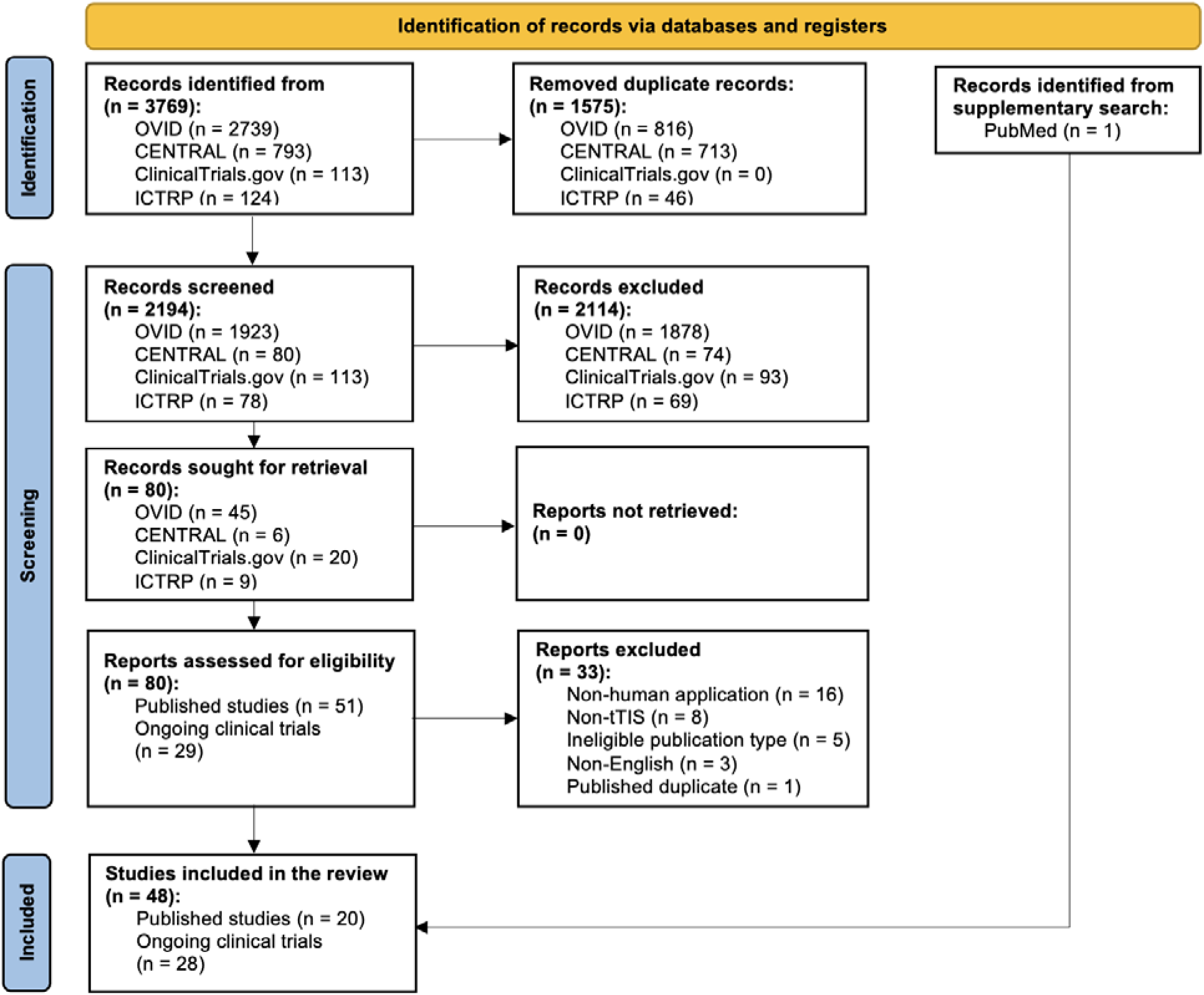
PRISMA flow diagram illustrating the study selection process for the systematic review examining tTIS applications in humans.

**Figure 2.**
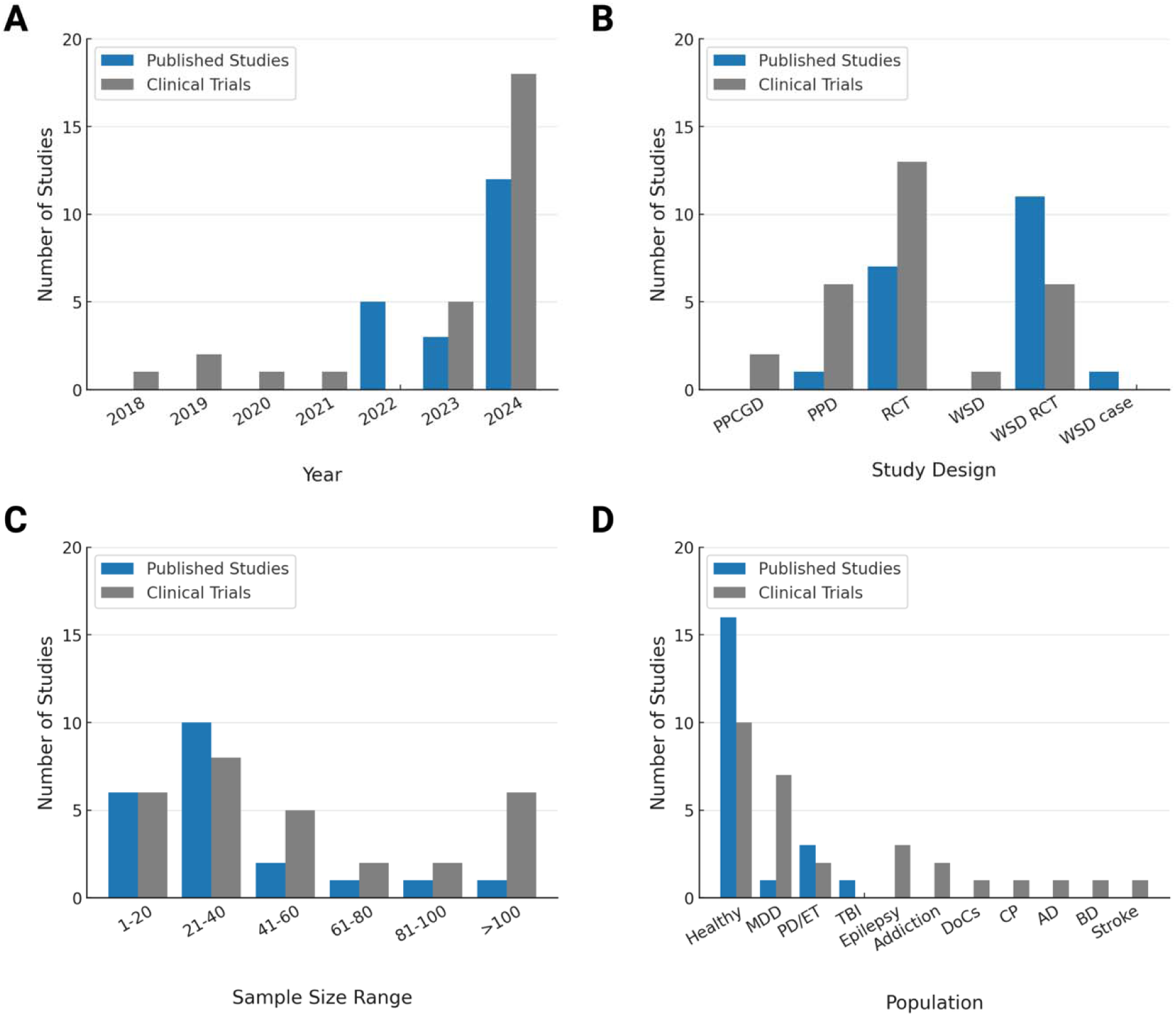
Trends, Study Designs, and Clinical Populations in Human tTIS Research. **(A)** Number of published studies and ongoing clinical trials involving human tTIS from 2018 and 2024. (**B)** Study design types among published human tTIS studies and ongoing clinical trials. (**C)** Sample size distribution in published human tTIS studies and ongoing clinical trials. (**D)** Study populations by condition in published human tTIS studies and ongoing clinical trials. **Abbreviations**: AD = Alzheimer’s disease; BD = Bipolar disorder; CP = Cerebral palsy; DoCs = Disorders of consciousness; ET = Essential tremor; MDD = Major depressive disorder; PD = Parkinson’s disease; PPCGD = Pretest-posttest control group design; PPD = Pretest-posttest design; RCT = Randomized controlled trial; TBI = Traumatic brain injury; WSD = Within-subjects design.

### 3.2. Research Design and Participants

Of the 20 published studies, 18 were randomized controlled trials (RCTs) [27–44], including 11 within-subjects crossover designs [27,29,31–35,38,41–43]; two used uncontrolled pretest-posttest designs [45,46] (**Figure 2B, eTable S4** in the **Supplement**). Fifteen involved healthy participants [27–36,38,39,41,42,44], one included both healthy and traumatic brain injury (TBI) participants [40], three studied Parkinson’s disease (PD) or essential tremor (ET) [43,45,46], and one published protocol [37] targets major depressive disorder (MDD) (**Figures 2C-D**). Sixteen studies were single-session [27–33,35,36,38,39,41–43,45,46]; four used multi-session protocols of 2-10 sessions (**eTable S5** in the **Supplement**) [34,37,40,44]. Sham (0 mA current) was used in 14 studies [27–30,32,33,36–41,43,44], tACS as an active control in 7 studies [29,30,32,34,39,40,42], and tDCS as a comparator in 2 studies [31,35].

Among the 28 ongoing trials, 16 use multi-session protocols with 2-40 sessions (**eTable S6** in the **Supplement**). In terms of scope, 16 feature mechanistic outcomes, while 19 focus on therapeutic applications of tTIS: 7 in MDD, 3 in epilepsy, 2 each in PD and addiction, and 1 each in Alzheimer’s disease, bipolar disorder, cerebral palsy, disorders of consciousness, or stroke (**Figure 2D**). Twenty-one ongoing trials are multi-arm: 6 feature different tTIS envelope frequency or target comparisons, 4 use tACS as an active control, 1 features tDCS as an active control, and 18 feature 0 mA stimulation as a sham control. Half of the ongoing tTIS trials (14 out of 28) are double-blind (**eTable S4** in the **Supplement**).

### 3.3. Methodological Quality of Studies

RCTs [27–44] showed moderate-to-high quality, with good baseline comparability of participants (16/18 studies) [27–35,37,38,40–44], intervention adherence (17/18 studies) [27–38,40–44], and outcome assessment (18/18 studies) [27–44] (**eTable S1** in the **Supplement**). The mean quality score was 10.2 ± 1.4 out of 14 (range: 8-14). Several studies lacked a detailed report of randomization methods (7/18 studies) [28,29,31,32,36,38,40], allocation concealment (13/18 studies) [27–29,31–34,36,38,40–43], and power calculations (9/18 studies) [30–32,36–38,41,42,44]. The two uncontrolled studies [45,46] had consistent intervention delivery, outcome assessment, and low attrition (2/2 studies) but moderate quality (mean score: 7.0 ± 0.0 out of 11), as they failed to fully address sample size adequacy and the use of statistics (**eTable S2** in the **Supplement**). **Figure 3** presents the status of published and ongoing tTIS human research, as well as allocation and blinding methods. China was the leading contributor to the field (**Figures 4A-B**), and most published work focused on safety (16 studies) [27–30,33,36–46] and behavioural outcomes (13 studies) [27–30,33,34,36–39,41,42,44] (**Figure 4C, eTables S4-6** in the **Supplement**).

**Figure 3.**
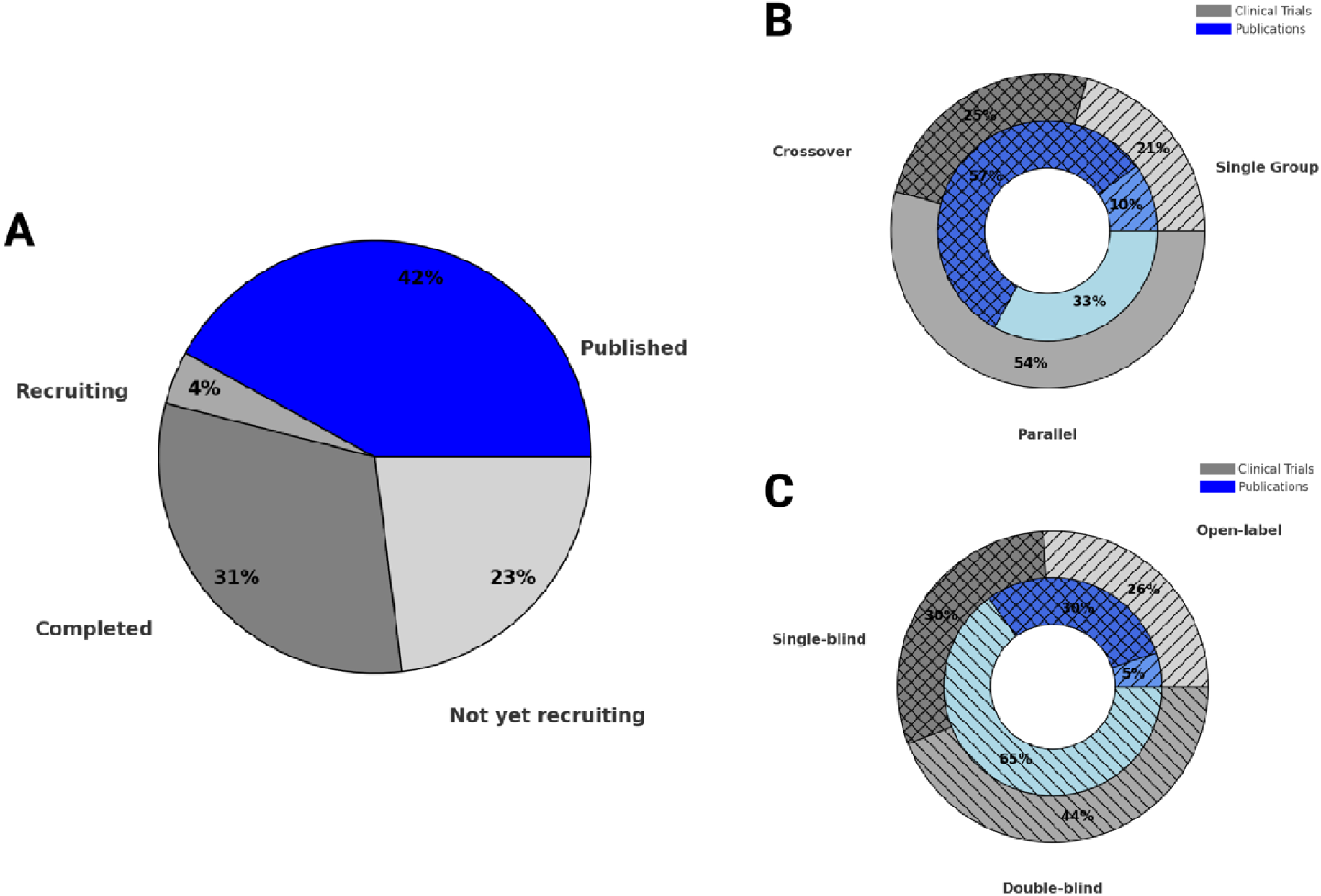
Status, Allocation, and Blinding Methods of Human tTIS Studies and Ongoing Clinical Trials. **(A)** Status of published human tTIS studies (completed) and ongoing clinical trials. **(B)** Distribution of group assignment methods in published human tTIS studies and ongoing clinical trials. **(C)** Distribution of masking methods used in published human tTIS studies and ongoing clinical trials.

**Figure 4.**
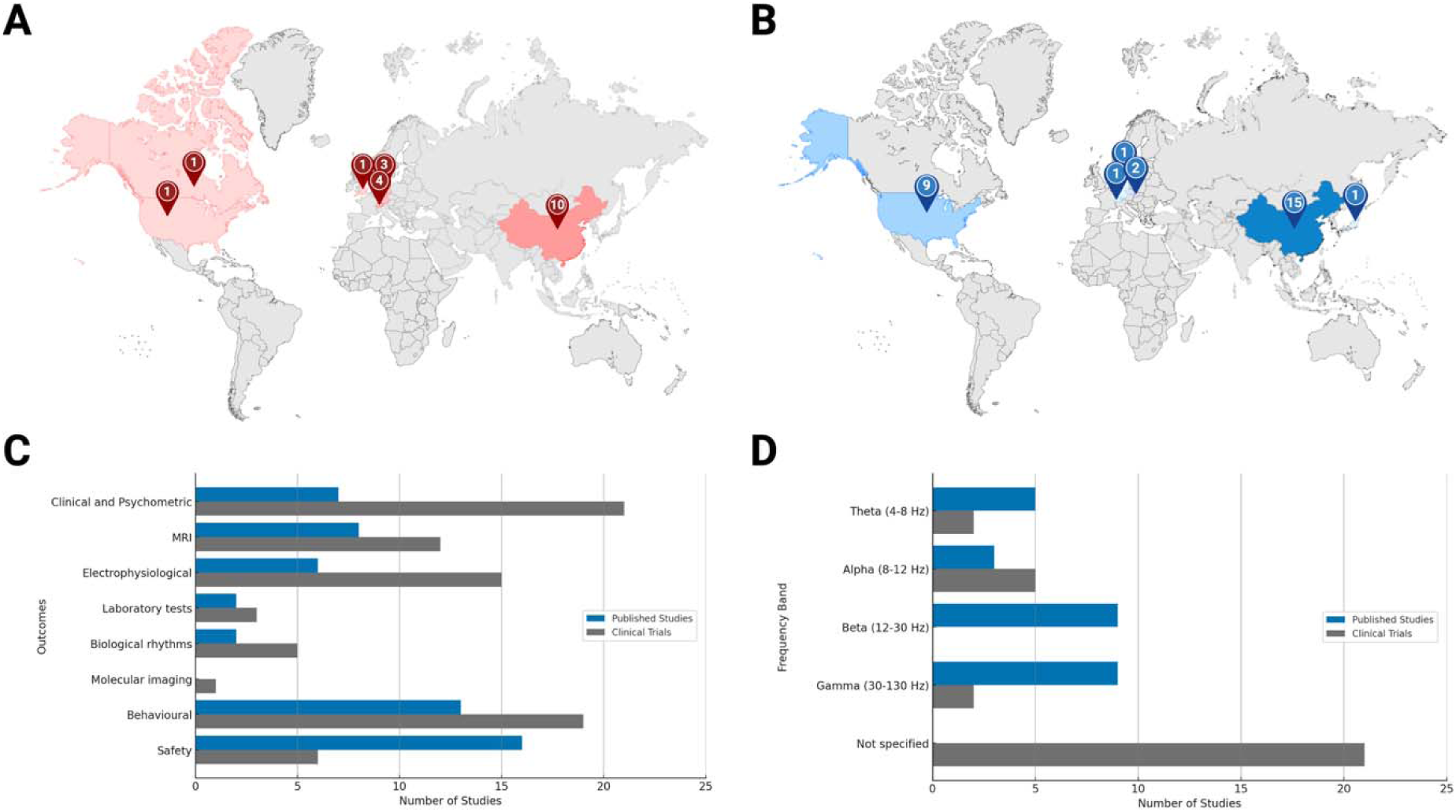
Global Landscape and Methodological Characteristics of Human tTIS Studies and Ongoing Clinical Trials. **(A)** Geographic distribution of published human tTIS studies, highlighting regions actively contributing to the field. *Map lines delineate study areas and do not necessarily depict accepted national boundaries*. **(B)** Geographic distribution of ongoing human tTIS clinical trials, reflecting current global research efforts. *Map lines delineate study areas and do not necessarily depict accepted national boundaries*. **(C)** Outcome measures reported in published human tTIS studies and those being collected in ongoing clinical trials. **(D)** tTIS envelope frequency bands used in published human tTIS studies and ongoing clinical trials, showing variation in stimulation parameters. **Abbreviations:** MRI = Magnetic resonance imaging.

### 3.4. Brain Targets and Stimulation Parameters

Envelope frequencies ranged from 5-130 Hz in published studies. Nine studies administered beta-range tTIS (20 Hz) [27,28,31,35,36,38,40,41,44], and nine used gamma-range tTIS (30-130 Hz) [27,28,36,37,40,41,43,45,46] (**Figure 4D, Table 1**). Carrier frequencies ranged from 0.90-2.13 kHz across all but one study, with 14 studies using pairs centred around 2 kHz [27,28,30–35,38,40–42,44,46]; one outlier study employed 20.00-20.07 kHz carriers [36]. Amplitudes ranged from 0.5-3 mA (zero-to-peak) in all studies except the same outlier, which used 15 mA; 17 studies applied 1-3 mA/channel with 5-30 s ramp-ins and 10-30 min of stimulation [27,28,30–35,37–44,46] (**eTable S7** in the **Supplement**). Reporting of impedance monitoring varied considerably across studies. Thirteen of twenty studies (65%) documented electrode-skin impedance testing [27–29,32–34,37–40,42,43,45], of which ten specified thresholds, most commonly maintaining values below 20 kΩ. Three studies described testing without providing thresholds [33,42,45], while seven studies (35%) did not report impedance monitoring at all [30,31,35,36,41,44,46] (**Table 1**).

**Table 1.** Summary of Stimulation Parameters in Human tTIS Studies.

| Study | Brain target | tTIS montage | tTIS waveform | Envelope frequency, $\Delta f$ | Carrier frequencies, $f_1/f_i + \Delta f$ | Zero-to-peak amplitude, $I_1 \vert I_2$ | Impedance | Ramp time | tTIS device manufacturer |
| --- | --- | --- | --- | --- | --- | --- | --- | --- | --- |
| Ma et al. (2022) | Left M1 | 30 mm away from M1 hotspot along FPZ-OZ & T3-T4 axes | Sinusoidal | 20 Hz<br>70 Hz | 2000 Hz / 2020 Hz<br>2000 Hz / 2070 Hz | 1 mA \| 1 mA | < 10 kΩ | 30 s | JUNTEX, Zhengzhou, China |
| Piao et al. (2022) | Left M1 | FC3-C5 & CP3-C1 | Sinusoidal | 20 Hz<br>70 Hz | 2000 Hz / 2020 Hz<br>2000 Hz / 2070 Hz | 1 mA \| 1 mA | < 10 kΩ | 30 s | Custom-built |
| von Conta et al. (2022) | Parieto-occipital cortex | C3-O1 & C4-O2 | Sinusoidal | IAF | (1000 – IAF/2) Hz \| (1000 + IAF/2) Hz | 0.5 mA \| 0.5 mA | < 20 kΩ | 10 s | NeuroConn GmbH, Ilmenau, Germany |
| Zhang et al. (2022) | dIPFC, IPL | 5 cm around F4 and P4 (4 pairs) | Sinusoidal | 6 Hz | 2000 Hz \| 2006 Hz | 1 mA \| 1 mA | ns | 15 s | Custom-built |
| Zhu et al. (2022) | Left M1 | M1 hotspot: A1-A2, A1-B1, B1-B2, A2-B2 | Sinusoidal | 20 Hz | 2000 Hz \| 2020 Hz | 1 mA \| 1 mA | ns | 30 s | Soterix Medical, NJ, USA |
| Iszak et al. (2023) | Occipital cortex | O1-C3 & O2-C4 | Sinusoidal | 10 Hz | 2000 Hz \| 2010 Hz | 1.65-2 mA \| 1.65-2 mA | < 22 kΩ | 25 s | NeuroConn GmbH, Ilmenau, Germany |
| Violante et al. (2023) | Left hippocampus | e<sub>1</sub>-e<sub>2</sub> and e<sub>3</sub>-e<sub>4</sub><br>e<sub>1</sub> and e<sub>3</sub> on the left hemisphere's nasion plane 5 cm apart, e<sub>2</sub> and e<sub>4</sub> above the right eyebrow 16 cm apart | Sinusoidal | 5 Hz | 2000 Hz \| 2005 Hz | 2 mA \| 2 mA<br>1 mA \| 3 mA | Tested, threshold ns | 5 s | Custom-built |
| Wessel et al. (2023) | Bilateral striatum | F3-F4 & TP7-TP8 | Sinusoidal | 100 Hz (iTBS) | 2000 Hz \| 2100 Hz every 10 s | 2 mA \| 2 mA | < 20 kΩ | 5 s | Digitimer, Letchworth Garden City, UK |
| Beanato et al. (2024) | Right hippocampus | P7-CP8 & FP1-FT8 | Sinusoidal | 100 Hz (cTBS; iTBS) | 2000 Hz \| 2100 Hz (iTBS: every 10 s) | 2 mA \| 2 mA | Tested, threshold ns | 5 s | Digitimer, Letchworth Garden City, UK |
| Demchenko et al. (2024) | Bilateral sgACC | AF7-T7 & AF8-T8 | Sinusoidal | 130 Hz | 1000 Hz \| 1130 Hz | 2 mA \| 2 mA | Adaptive (target: < 25 kΩ) | 30 s | Soterix Medical, NJ, USA |
| Liu et al. (2024) | Bilateral SN | F5-P5 & F6-PO8 | Sinusoidal | 130 Hz | 900 Hz \| 1030 Hz | 0.75-1 mA \| 0.75-1 mA | Tested, threshold ns | ns | Jiangsu Jinyuan Medical Technology Co., Xuzhou, Jiangsu, China |
| Modak et al. (2024) | Left caudate | F9-F10 & FP1-CPZ | Sinusoidal | 20 Hz | 2000 Hz \| 2020 Hz | 2 mA \| 2 mA | < 50 kΩ | 30 s | Soterix Medical, NJ, USA |
| Thiele et al. (2024) | Parieto-occipital cortex | P4-I1/O1 & P3-I2/O2<sup>a</sup> | Sinusoidal | IAF | 1000 Hz \| 1000 + IAF Hz | 1 mA \| 1 mA | < 5 kΩ | 10 s | NeuroConn GmbH, Ilmenau, Germany |
| Vassiliadis et al. (2024a)<sup>b</sup> | Bilateral striatum | F3-F4 & TP7-TP8 | Sinusoidal | 20 Hz<br>80 Hz | 1990 Hz \| 2010 Hz<br>1960 Hz \| 2040 Hz | 2 mA \| 2 mA | ns | 5 s | Digitimer, Letchworth Garden City, UK |
| Vassiliadis et al. (2024b)<sup>c</sup> | Bilateral striatum; left hippocampus | F3-F4 & TP7-TP8; P8-CP7 & FP2-FT7 | Sinusoidal | 100 Hz (cTBS, iTBS)<br>20 Hz<br>80 Hz | 2000 Hz \| 2100 Hz (iTBS: every 10 s)<br>1990 Hz \| 2010 Hz<br>1960 Hz \| 2040 Hz | 0.5-2 mA \| 0.5-2 mA | < 20 kΩ | 5 s | Digitimer, Letchworth Garden City, UK |
| Wang et al. (2024) | Left M1 | 3 cm away from C3 (2 pairs) | Sinusoidal | 20 Hz<br>70 Hz | 20000 Hz \| 20020 Hz<br>20000 Hz \| 20070 Hz | 15 mA \| 15 mA | ns | 30 s | Custom-built |
| Yang et al. (2024a)<sup>d</sup> | Right GPi | CP3-CP6 & F3-F6 | Sinusoidal | 130 Hz | 1300 Hz \| 1430 Hz | 2.5 mA \| 2 mA | < 15 kΩ | 30 s | Soterix Medical, NJ, USA |
| Yang et al. (2024b)<sup>e</sup> | Contralateral STN | Individualized | Sinusoidal | 130 Hz | 2000 Hz \| 2130 Hz | 1.5-2 mA \| 1.5-2 mA | ns | 30 s | NeuroDome Medical Technology Co., Xi'an, Shaanxi, China |
| Zheng et al. (2024) | Bilateral M1 (leg area) | F3-P3 & F4-P4 | Sinusoidal | 20 Hz | 2000 Hz \| 2020 Hz | 1 mA \| 1 mA | ns | 30 s | National Instruments, TX, USA<br>World Precision Instruments, FL, USA |
| Zhu et al. (2024) | Left M1 | M1 hotspot: A1-A2, A1-B1, B1-B2, A2-B2 | Sinusoidal | 20 Hz | 2000 Hz \| 2020 Hz | 1 mA \| 1 mA | ns | 30 s | Soterix Medical, NJ, USA |

**Abbreviations:** cTBS = Continuous theta burst stimulation; dlPFC = Dorsolateral prefrontal cortex; $\Delta f$ = Envelope frequency; $f_l$ = Carrier frequency; GPi = Globus pallidus internus; IAF = Individual alpha frequency; $I_1$ , $I_2$ = Current intensity; iTBS = Intermittent theta burst stimulation; IPL = Inferior parietal lobule; M1 = Primary motor cortex; ns = Not specified; sgACC = Subgenual anterior cingulate cortex; SN = Substantia nigra; STN = Subthalamic nucleus.
cTBS: bursts of 3 pulses at 100 Hz delivered at 5 Hz
iTBS: bursts of 3 pulses at 100 Hz repeated at 5 Hz for 2 s, interspersed with 8 s without any stimulation
<sup>a</sup>The second electrode in the electrode pair was positioned between I1 and O1 for channel 1, and between I2 and O2 for channel 2.
<sup>b</sup>Vassiliadis et al. (2024). *Nat Hum Behav.*
<sup>c</sup>Vassiliadis et al. (2024). *J Neural Eng.*
<sup>d</sup>Yang et al. (2024). *Mov Disord.*
<sup>e</sup>Yang et al. (2024). *Brain Stimul.*

Common targets for tTIS were the primary motor cortex (M1) [27,28,31,35,36,44], parieto-occipital cortex [29,32,39], basal ganglia [34,38,40,41,43,45,46], and hippocampus [33,40,42] (**Figure 5**). Theta-range tTIS was typically used for the hippocampus [33,40,42] and striatum [34,40]; alpha-range for parieto-occipital cortex [29,32,39]; beta-range for M1 and striatum [27,28,31,35,36,38,40,41,44]; 130 Hz for basal ganglia and subgenual anterior cingulate cortex [37,43,45,46]. Emerging trials often use 10 Hz or 130 Hz envelopes targeting brain regions such as the dorsolateral prefrontal cortex, amygdala, and basal ganglia structures. Across ongoing trials, carrier frequencies cluster in the 1.30-2.01 kHz range, with envelope frequencies tailored to the clinical indication (e.g., 20 Hz for motor or PD, theta/alpha for MDD, gamma for mechanistic motor studies). Peak intensities vary from 0.85-4.36 mA (zero-to-peak), and session durations range from 20-60 min (**eTable S8** in the **Supplement**). Cumulative exposure spans 2-40 sessions depending on protocol design.

**Figure 5.**
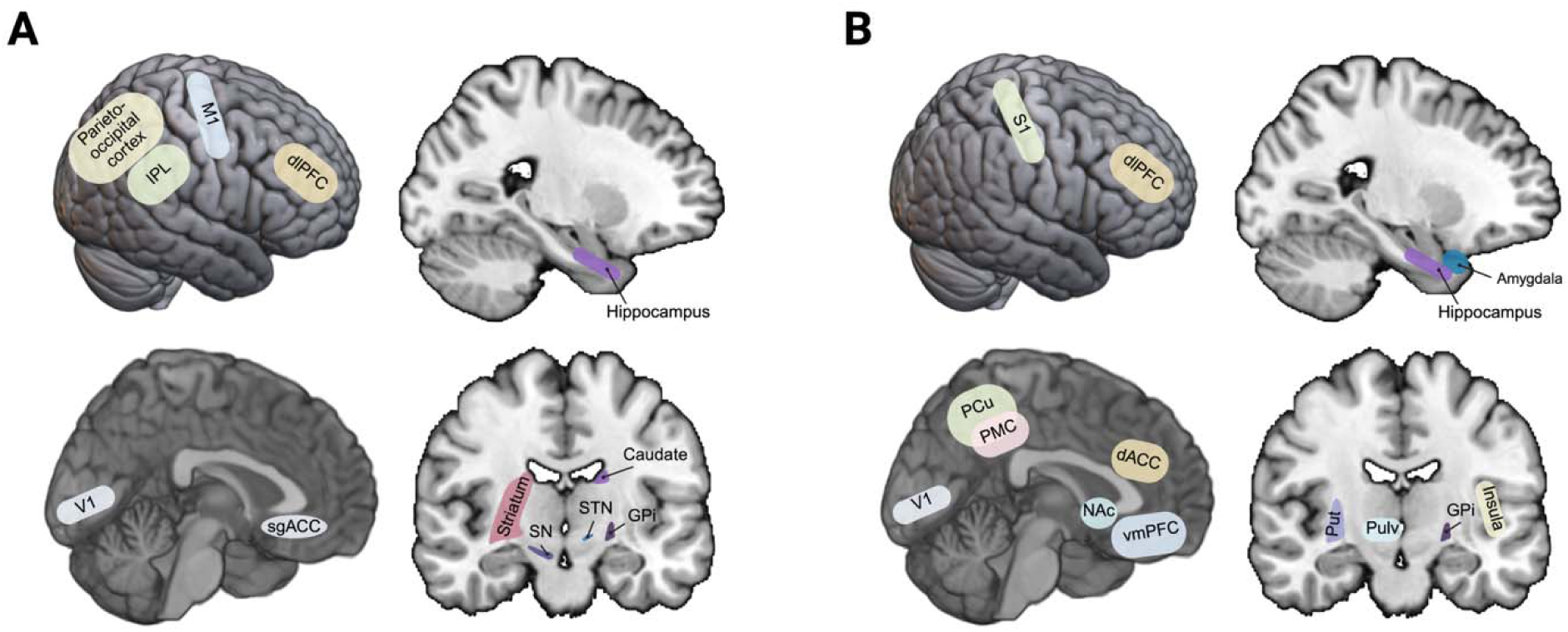
Brain Targets in Human tTIS Studies and Ongoing Clinical Trials. **(A)** Brain regions targeted in published human tTIS studies, illustrating early applications of the technique. **(B)** Brain regions targeted in ongoing human tTIS clinical trials, reflecting current translational priorities and therapeutic goals. **Abbreviations:** dACC = Dorsal anterior cingulate cortex; dlPFC = Dorsolateral prefrontal cortex; GPi = Globus pallidus internus; IPL = Inferior parietal lobule; M1 = Primary motor cortex; NAc = Nucleus accumbens; PCu = Precuneus; PMC = Posteromedial cortex; Pulv = Pulvinar nuclei; Put = Putamen; S1 = Primary somatosensory cortex; sgACC = Subgenual anterior cingulate cortex; SN = Substantia nigra; STN = Subthalamic nucleus; V1 = Primary visual cortex; vmPFC = Ventromedial prefrontal cortex.

### 3.5. Safety and Tolerability Outcomes

Sixteen studies reported safety and tolerability outcomes [27–30,33,36–46]. In 10 studies differentiating AEs between tTIS and control groups [27,28,30,33,36,40,41,43,45,46], tTIS was associated with higher rates of tingling (*P* < .05) and itching (*P* < .001), with no serious AEs or epileptic activity (**Table 2**). Sensation ratings were comparable between tTIS and control groups [33,38,40], although older adults reported reduced intensity [40]. Across all studies, only one TBI participant withdrew due to strong sensations [40]. Among ongoing trials, 6/28 (21%) explicitly report plans to collect safety data.

**Table 2.** Pooled Frequencies of Adverse Events Reported in Human tTIS Studies.

| Adverse Event, *n* (%) | tTIS<br>*n* = 333 | Control<br>*n* = 282 | $\chi^2$ | *P* Value |
| --- | --- | --- | --- | --- |
| Tingling | 35 (11) | 13 (5) | 6.59 | *P* < **0.05** |
| Itching<sup>a</sup> | 31 (11) | 9 (3) | 11.95 | *P* < **0.001** |
| Warmth | 18 (5) | 6 (2) | 3.54 | *P* = 0.06 |
| Burning | 8 (2) | 4 (1) | 0.34 | *P* = 0.05 |
| Headache | 18 (5) | 10 (4) | 0.83 | *P* = 0.36 |
| Fatigue<sup>b</sup> | 38 (13) | 24 (10) | 0.87 | *P* = 0.35 |
| Sleepiness/drowsiness | 9 (3) | 8 (3) | $6.06 \times 10^{-29}$ | *P* > 0.99 |
| Dizziness/vertigo | 13 (4) | 9 (3) | 0.07 | *P* = 0.80 |
| Nausea | 5 (2) | 2 (<1) | 0.29 | *P* = 0.59 |
| Pain | 6 (2) | 6 (2) | $2.11 \times 10^{-32}$ | *P* > 0.99 |

<sup>a</sup> *n* (tTIS) = 281 participants, *n* (Control) = 282 participants. Yang et al. (2024a) excluded due to non-differentiation of itching from general discomfort.
<sup>b</sup> *n* (tTIS) = 283 participants, *n* (Control) = 232 participants. Ma et al. (2022) excluded due to an unclear number of participants reporting fatigue across sessions.

### 3.6. Clinical Outcomes

Three studies evaluated the clinical effects of 130 Hz tTIS targeting the globus pallidus internus (GPi) [43], subthalamic nucleus (STN) [46], and substantial nigra (SN) in PD and ET [45] (**eTable S9** in the **Supplement**). A double-blind RCT [43] targeting the GPi showed 14.7% reduction in overall symptom severity based on the Movement Disorder Society-Unified Parkinson’s Disease Rating Scale Part III [47] (MDS-UPDRS-III; *P* = .02), with significant improvements in bradykinesia (23.5%, *P* = .01) and tremor (15.3%, *P* = .01). A case series [45] involving 2 PD and 1 ET patients reported reduced tremor amplitude with tTIS over SN compared to tACS control. An open-label trial [46] targeting the STN showed 27.5% symptom reduction on MDS-UPDRS-III immediately post-stimulation, with moderate-to-large effect sizes for overall symptom severity (Hedge’s *g* = −0.92), bradykinesia (Hedge’s *g* = −0.72), and rigidity (Hedge’s *g* = −0.88), and smaller effect sizes for tremor (Hedge’s *g* = −0.35) and axial symptoms (Hedge’s *g* = −0.28) (**eTable S10** in the **Supplement**). Among ongoing trials, 21/28 (75%) plan to assess clinical or psychometric outcomes.

### 3.7. Behavioural Outcomes

Of 13 behavioural studies [27–30,33,34,36–39,41,42,44], six investigated motor function with tTIS targeting M1 [27,28,36,44] or striatum [34,41]. 20 Hz tTIS over M1 showed mixed results: no change in reaction time (RT), dexterity, or postural stability [27,28,36,44], but improved implicit motor learning [27] and vertical jump performance [44] (**eTable S11** in the **Supplement**). Striatal studies showed frequency-dependent effects: 100 Hz intermittent theta-burst stimulation (iTBS)-patterned tTIS enhanced motor learning gains in a sequential finger tapping task [34], while 80 Hz disrupted reinforcement-related motor learning [41].

Five studies assessed tTIS effects on memory and cognition [29,30,33,38,42]. Working memory (WM) showed minimal to no improvement following tTIS [29,30,38]. In contrast, hippocampal tTIS improved spatial navigation efficiency [42] and episodic face-name recall [33]. Two visual studies showed no effects on mental rotation or phosphene induction [32,39]. Among ongoing trials, 19/28 (68%) plan to assess behavioural outcomes.

### 3.8. Neuroimaging Outcomes

Seven studies used functional magnetic resonance imaging (fMRI) to validate target engagement following tTIS [31,33–35,38,41,42]. Two M1 studies reported medium-to-large increases in resting-state activity and functional connectivity (FC) within sensorimotor networks [31,35] (**eTable S12** in the **Supplement**). Hippocampal tTIS produced medium effects, reducing activity evoked by memory tasks, decreasing FC within the anterior-temporal network, and disrupting spatial coding [33,42]. Three striatal studies [34,38,41] demonstrated medium-to-large target activation and strengthened striatal-frontal FC during reinforcement learning, suggesting plasticity effects during active learning but not at rest. Among ongoing trials, 12/28 (43%) plan to collect MRI data.

### 3.9. Neurophysiological Outcomes

Five studies investigated the neurophysiological effects of tTIS [28,29,32,36,39]. Alpha-range tTIS over parieto-occipital cortex yielded null-to-small effects on resting alpha power in three studies [29,32,39]; one of these [39], however, reported a medium effect with increased alpha event-related desynchronization during a mental rotation task (**eTable S12** in the **Supplement**). Two M1 studies [28,36] applying beta-(20 Hz) or gamma-range (70 Hz) tTIS showed null effects on electroencephalography (EEG) band power. Among ongoing trials, 15/28 (54%) include neurophysiological measures, and 5/28 (18%) will collect outcomes related to sleep or fatigue.

## 4. Discussion

This systematic review summarizes emerging trends in human tTIS research. While most studies to date have focused on safety and mechanistic outcomes in healthy participants and can thus be considered Phase 1 trials, preliminary clinical investigations, particularly in PD and ET, suggest that tTIS may offer acute motor symptom improvement. Across studies, tTIS was most commonly delivered at beta or gamma envelope frequencies to modulate neural oscillations implicated in motor control and cognitive functions [48,49]. Neuroimaging findings support target engagement of the M1, basal ganglia, and hippocampus with tTIS, highlighting its potential as a non-invasive tool for targeted neuromodulation in humans.

Our review revealed several methodological trends that may guide future research. The most widely adopted protocol involved carrier frequencies centred around 2 kHz, envelope frequencies of 20 Hz (beta range), and current amplitudes of 1-3 mA per channel, delivered for 10-30 min with ramp-ups of 5-30 s. These parameters likely reflect a balance between tolerability, field strength, and theoretical considerations of minimizing unintended neuronal activation. Frequency choices were generally informed by prior knowledge of region-specific oscillatory dynamics: theta-range envelopes were most often applied to the hippocampus and striatum, in line with the role of theta oscillations in memory encoding and fronto-striatal communication [50,51]; alpha-range to the parieto-occipital cortex, consistent with alpha’s role in visual attention and sensory gating [52]; beta-range to M1 and striatum, reflecting beta’s involvement in motor control and basal ganglia-cortical interactions [53]; and 130 Hz to basal ganglia and subgenual anterior cingulate cortex, paralleling high-frequency DBS paradigms in PD and treatment-resistant depression [54,55]. Nevertheless, as the sufficiency of 1 kHz carriers was uncertain in the earlier phases of tTIS research in humans, some studies employed ∼1 kHz while others investigated ∼2 kHz, reflecting ongoing efforts to establish optimal stimulation parameters. Carrier frequencies should be set to ≥2 kHz in order to minimize off-target neuronal activation [56–58]. Current amplitudes generally ranged from 0.5-2.5 mA, well below published safety thresholds. Although theoretical guidelines permit up to ∼16 mA at carrier frequencies below 2.5 kHz [59,60], practical considerations such as brain heating (∼14 mA) and cutaneous stimulation (∼7 mA) impose lower limits. One notable exception was Wang et al. (2024) [36], who applied 15 mA at 20 kHz. While this protocol remains within frequency-dependent safety thresholds, as Cassarà et al. (2025) [59,60] describe allowable current as increasing linearly with frequency above 2.5 kHz, it relied primarily on phantom-based current density estimates and did not report impedance or voltage monitoring, underscoring the need for comprehensive in vivo safety monitoring and standardized reporting in future trials. Although our review focuses on transcranial applications, it is also worth noting that interferential stimulation in peripheral muscle contexts employs substantially higher amplitudes and carrier frequencies without eliciting cutaneous discomfort, highlighting potential differences in tolerability across temporal interference stimulation modalities.

Safety outcomes were favourable across studies, with generally mild AEs (e.g., tingling, itching) and no serious AEs. These findings are in line with the largest human tTIS safety investigation to date [40], suggesting the overall tolerability of tTIS in humans. However, variability in impedance monitoring practices highlights an important methodological gap, with only 65% of studies reporting electrode-skin impedance testing. Future trials should implement and report standardized impedance monitoring protocols throughout stimulation sessions, using established thresholds to enable more comprehensive safety assessment and facilitate cross-study comparisons. Of note, as a function of its high-frequency carriers [36], tTIS generally elicits less cutaneous sensation than tACS despite requiring higher currents for effective neuromodulation. Because sensations are minimal and less distinguishable from sham, this may improve blinding efficiency relative to conventional tACS [33,61], but also reduces the likelihood of early subjective warning signs if unsafe stimulation occurs in the absence of adequate monitoring.

Second, only four studies to date have employed multi-session tTIS protocols [34,37,40,44], although several upcoming trials plan to incorporate up to 40 sessions. This shift, alongside the expanding application of tTIS in various neurological and psychiatric disorders, reflects growing interest in the therapeutic potential of multi-session tTIS. Control conditions, however, remain variable across studies, underscoring the need for methodological standardization and rigorous blinding. While sham stimulation with no current (0 mA) is commonly used, it may not sufficiently account for sensory confounds associated with high-frequency carrier exposure. A more appropriate alternative is an active control condition in which two high-frequency alternating currents are applied without a frequency difference, thereby eliminating the low-frequency interference envelope while preserving comparable scalp sensations [62]. Since tTIS is a specialized form of tACS that uses two out-of-phase high-frequency currents, multi-channel tACS may more closely approximate its sensory and physiological effects. This could, in principle, make it a more mechanistically appropriate control than no-current sham or conventional single-channel tACS, although additional assumptions are needed to determine whether it truly allows for better isolation of the specific effects of the interference pattern.

Clinical translation of tTIS also raises several practical challenges that warrant careful consideration. First, electric field modelling methods vary substantially across groups, and differences in head tissue conductivity values, anatomical fidelity, or mesh resolution can markedly alter estimates of focality and field strength, potentially impacting protocol reproducibility [17,63–65]. Standardization of modelling approaches and transparent reporting of conductivity assumptions will therefore be essential. Second, although most published studies report monitoring of electrode impedance, systematic safety monitoring remains inconsistent, and should include standardized assessment of voltage burden, skin temperature, and AE tracking across trials [66–68]. Third, it remains an open question whether stimulation parameter limits established in healthy volunteers are directly applicable to patient populations, particularly in individuals with neurological or psychiatric disorders who may exhibit altered tissue properties, sensitivity to stimulation, or a higher risk of AEs [69,70]. Establishing evidence-based distinctions in stimulation limits for patients versus healthy individuals will be an important step for future guideline development.

Clinically, tTIS has primarily been explored in PD and ET [43,45,46], while its safety has also been demonstrated in individuals with TBI [40]. Motor improvements have been reported following the stimulation of basal ganglia targets in patients with PD and ET [43,45,46]. The greatest improvements were observed in rigidity and bradykinesia, consistent with effects seen in unilateral STN DBS [71]. The open-label trial [46] also reported a larger decrease in MDS-UPDRS-III scores compared to the RCT [43] (27.5% vs 14.7%), likely due to differences in study design (uncontrolled vs. sham-controlled; medication-OFF vs. medication-ON) and stimulation target (STN vs. GPi). Of note, the RCT [43] targeting the right GPi reported significant improvements, particularly in contralateral motor function, consistent with the anatomy of motor control pathways [72]. In contrast, the open-label trial [46] observed stronger ipsilateral effects from unilateral STN stimulation, which may reflect cross-hemispheric connectivity within basal ganglia networks [71,73]. However, these results should be interpreted with caution, as the apparent asymmetry in motor improvement could also be influenced by the inherent lateralization of Parkinson’s disease symptoms, where the more affected limb often shows higher baseline impairment [74]. Both studies included small samples (8-15 patients) and assessed outcomes only up to 60 minutes after a single tTIS session, with no long-term follow-up data available. While these early findings suggest that tTIS can modulate motor circuits in movement disorders, larger RCTs are needed to determine whether these acute effects are reproducible and sustained over multiple sessions and longer follow-up periods. Importantly, the ongoing clinical trial landscape reflects this shift: nearly two-thirds of registered trials now focus on therapeutic indications, with multi-session protocols (2-40 sessions) and double-blind randomized designs increasingly common. Collectively, these trials may help establish whether the preliminary efficacy signals observed to date generalize across conditions and whether tTIS can achieve clinically meaningful and durable outcomes beyond acute mechanistic effects.

Interestingly, bradykinesia and tremor have consistently emerged as symptoms showing potential benefits with tTIS targeting the basal ganglia. A case series [45] also reported reductions in resting tremor following bilateral SN stimulation in three patients, raising the possibility that tTIS may replicate some effects of DBS by modulating pathological beta oscillations (12–30 Hz) [75–77]. The frequent use of 130 Hz envelopes mirrors conventional DBS protocols [78] and aligns with preclinical findings [79,80] of beta-range tTIS enhancing synaptic strength and plasticity in rodent motor circuits. However, mechanistic evidence in humans remains limited, and future studies incorporating EEG, fMRI, or invasive recordings of local field potentials through new DBS systems [81] should be considered.

In healthy populations, tTIS showed modest effects on motor outcomes, although some studies report frequency-dependent improvements in jump performance [44] or motor learning [27,34,41]. tTIS may help counteract age-related plasticity declines, based on findings [34] that striatal 100 Hz iTBS-patterned tTIS—approximating the lower therapeutic range of DBS [82]— accelerated motor adaptation in older adults. Replication of such findings is needed, and to determine their therapeutic value, future RCTs may consider evaluating tTIS as an adjunct to motor rehabilitation in aging populations or individuals with motor impairment.

Cognitive findings, on the other hand, remain variable. WM effects of tTIS remain limited [29,38], although some studies reported subtle improvements [30]. Hippocampal-targeted tTIS has shown promise in enhancing spatial navigation and episodic memory [33,42], particularly when stimulation was aligned with task-relevant timing and frequency. A key challenge in cognitive applications of tTIS is selecting behavioural tasks that accurately probe the function of targeted circuits [83,84]; such paradigms should be both sensitive and anatomically specific. As with other brain stimulation methods [85,86], tTIS appears to be more effective when the targeted network is actively engaged during stimulation rather than during the resting state, highlighting the importance of task-stimulation coupling to enhance both neural and behavioural effects [34].

One of the main advantages of tTIS is its potential for spatially selective targeting with minimized off-target effects [13,17,87,88]. Functional neuroimaging evidence supports this specificity, demonstrating successful neuromodulation of the motor [31,35], striatal [34,38,41], and hippocampal [33,42] circuits in humans. For instance, fMRI revealed striatal tTIS effects in the putamen, correlating with improved motor task performance [34]. Hippocampal tTIS using theta-range offsets reduced blood-oxygen-level-dependent (BOLD) signals during memory tasks and altered entorhinal activity [33,42], suggesting network-specific engagement. Region- and frequency-specific effects were also observed: 20 Hz tTIS over left M1 reduced dynamic FC variability yet increased mean FC strength within the sensorimotor network [35], while hippocampal theta-range tTIS modulated subregion-specific FC depending on current amplitude ratios between stimulation channels (1:3 vs. 1:1 mA) [33]. Specifically, the 1:3 montage reduced FC in the middle and posterior hippocampal subregions, whereas the 1:1 montage primarily modulated anterior and middle subregions [33]. These findings highlight how stimulation parameters shape regional specificity, suggesting that tailoring envelope frequencies and amplitudes, along with optimizing electrode montages using individualized computational models [88,89], may enhance focality. Closed-loop protocols [90,91] may further improve precision and efficacy, maximizing target engagement.

Nevertheless, despite growing evidence of motor and some cognitive benefits, the ability of tTIS to reliably modulate brain oscillations remains inconsistent. Alpha-range tTIS has yielded no effects on resting alpha power [28,29,32], with some evidence for task-related modulation [39]. Beta- and gamma-range tTIS over M1 showed no EEG effects in two studies [28,36], suggesting that tTIS may be more effective in modulating task-specific rather than resting-state neural dynamics, depending on protocol design, task choice, and level of behavioural engagement among study participants. Given high neural activation thresholds and tissue inhomogeneity, eliciting robust deep intracranial effects with low-intensity transcranial currents remains challenging [92,93]. Future work may explore alternative non-sinusoidal waveforms (e.g., pulse-width modulated tTIS [94]) or higher intensities to improve neural entrainment.

## 5. Strengths and Limitations

Strengths of this review include its comprehensive coverage of both clinical and basic tTIS studies, detailed consideration of ongoing clinical trials, and critical synthesis of methodological, safety, therapeutic, and mechanistic dimensions. Limitations include the small number of RCTs, publication bias, and heterogeneity in stimulation protocols, which collectively constrain reproducibility and generalizability. Of these, insufficient sample sizes likely represent the most immediate barrier to external validity. For example, several mechanistic [32,38] and clinical [43,45,46] studies recruited fewer than 20 participants, which limits statistical power and increases susceptibility to bias. In contrast, heterogeneity in stimulation parameters, illustrated by the variation in envelope frequencies and electrode montages (for example, 20 Hz vs. 70 Hz in [27] and 20 Hz vs. 80 Hz in [41]), primarily hampers reproducibility and cross-study comparison rather than internal validity. Publication bias further complicates interpretation by favouring positive findings and limiting the visibility of null results, which may lead to inflated expectations of efficacy.

## 6. Future Directions

Future research should prioritize well-designed RCTs, multimodal mechanistic validation of tTIS effects, and systematic optimization of stimulation parameters, particularly under task engagement, to advance tTIS from experimental technique to clinically viable intervention. Adequately powered trials with a priori sample size estimation are needed to reduce the risk of spurious findings and improve reliability [95]. Consensus on reporting standards, including electrode placement, current amplitudes, carrier and envelope frequencies, and safety monitoring procedures, will be critical to enhance reproducibility and comparability across studies [66,96]. Finally, standardization of control conditions, such as matched-frequency tACS, may provide a more rigorous comparator until optimized sham paradigms are established, although this approach relies on additional assumptions regarding mechanisms of action and requires further empirical validation [13,97].

## 7. Conclusions

Preliminary Phase 1 studies demonstrate the safety, tolerability, and short-term clinical benefits of tTIS in PD and ET, with evidence of target engagement of motor, striatal, and hippocampal circuits across healthy and clinical populations. However, existing evidence is limited by small sample sizes and a lack of follow-up data, limiting conclusions about its therapeutic potential. Phase 2 trials are now needed to gather initial clinical efficacy data in patient populations, explore the effects of multi-session protocols, and assess the durability of effects. These trials should ideally use matched-frequency tACS as a control condition, with both channels delivering frequencies identical to the tTIS carrier frequency. This approach controls for peripheral sensations and non-specific effects, thereby isolating the interference mechanism unique to tTIS. Additionally, tTIS should be paired with carefully designed behavioural tasks tailored to the targeted neural circuits to maximize therapeutic specificity.

## Data Availability

ID and VB have full access to all the data in the manuscript and take responsibility for the integrity of the data and the accuracy of the data analysis.

## Glossary

Beta Oscillations: neural oscillations in the 13–30 Hz frequency range, commonly associated with motor control, attention, and certain cognitive functions.
Carrier Frequency: the high-frequency (typically kilohertz range) sinusoidal currents used in temporal interference stimulation to generate a modulating interference pattern. Neurons do not respond directly to these high frequencies.
Envelope Frequency: the low-frequency amplitude modulation (e.g., 10–130 Hz) resulting from the interference between two slightly different carrier frequencies in temporal interference stimulation. This frequency is within the range neurons can respond to.
Event-Related Desynchronization (ERD): a decrease in the power of specific EEG frequency bands, such as alpha or beta, during cognitive or motor tasks, indicating cortical activation.
Sham Stimulation: a placebo condition in neuromodulation studies in which no current is delivered (or a brief mimic current is applied) to blind participants and control for expectancy effects.
Temporal Interference Stimulation (tTIS): a non-invasive brain stimulation method that applies two high-frequency alternating currents with a slight frequency difference to create a low-frequency envelope at a specific deep brain target, enabling modulation of deep structures with minimal off-target effects.
Transcranial Alternating Current Stimulation (tACS): a neuromodulation technique that delivers sinusoidal alternating current through scalp electrodes to entrain or modulate brain oscillations at specific frequencies.
Transcranial Direct Current Stimulation (tDCS): a technique that applies a constant, low-intensity direct current through electrodes on the scalp to alter cortical excitability and promote plasticity.
Transcranial Magnetic Stimulation (TMS): a non-invasive brain stimulation technique that uses magnetic fields to induce electric currents in specific areas of the brain, widely used for research and clinical treatment of depression and other conditions.

## Acknowledgements

The authors thank Yu Liu for providing feedback on the manuscript.

## Data Availability

Data supporting this systematic review are available from the corresponding author upon reasonable request.

## CRediT Authorship Statement

**Ilya Demchenko**: Conceptualization, Data curation, Formal analysis, Investigation, Methodology, Project administration, Resources, Validation, Visualization, Writing – original draft, Writing – review and editing; **Ishaan Tailor**: Data curation, Formal analysis, Investigation, Methodology, Software, Validation, Writing – original draft, Writing – review and editing; **Sina Chegini:** Data curation, Formal analysis, Investigation, Methodology, Software, Validation, Visualization, Writing – original draft, Writing – review and editing; **Haochen Yu**: Data curation, Formal analysis, Investigation, Methodology, Software, Validation, Visualization, Writing – original draft, Writing – review and editing; **Fatemeh Gholamali Nezhad**: Investigation, Resources, Validation, Writing – original draft, Writing – review and editing; **Alice Rueda**: Resources, Validation, Writing – review and editing; **Anne Kever:** Resources, Validation, Writing – review and editing; **Sridhar Krishnan**: Resources, Validation, Writing – review and editing; **Abhishek Datta**: Resources, Validation, Writing – review and editing; **Jed A. Meltzer**: Resources, Supervision, Validation, Writing – review and editing; **Simon J. Graham**: Resources, Supervision, Validation, Writing – review and editing; **Tom A. Schweizer**: Resources, Supervision, Validation, Writing – review and editing; **Sumientra Rampersad**: Resources, Validation, Writing – review and editing; **Edward S. Boyden**: Resources, Validation, Writing – review and editing; **Ines R. Violante**: Resources, Validation, Writing – review and editing; **Robert Chen**: Resources, Validation, Writing – review and editing; **Andres M. Lozano**: Resources, Validation, Writing – review and editing; **Venkat Bhat**: Conceptualization, Investigation, Project administration, Resources, Supervision, Validation, Writing – review and editing.

