## Supplement for "Human Applications of Transcranial Temporal Interference Stimulation: A Systematic Review"

### eMethods

##### Search Strategy

###### OVID Databases Search

**Embase Classic+Embase** 1947 to 2024 December 11**, APA PsycInfo** 1806 to December 2024 Week 1**, Ovid**

**MEDLINE(R) ALL** 1946 to December 11, 2024

| **#** | **Searches** | **Results** |
| --- | --- | --- |
| 1 | exp Brain/ | **3525306** |
| 2 | exp Models, Neurological/ or exp computer model/ or exp computational modeling/ | **2603873** |
| 3 | exp Clinical Trial/ or exp Clinical Trials/ | **3475171** |
| 4 | exp Electric Stimulation Therapy/ or exp electrostimulation/ or exp electrostimulation therapy/ or exp electrotherapy/ or exp Electrical Stimulation/ or exp Brain Stimulation/ or exp Electrical Brain | **682700** |
|  | Stimulation/ |  |
| 5 | (clinical adj3 (stud* or trial* or project*)).tw,kf,id. | **2276457** |
| 6 | brain*.tw,kf,id. | **3600887** |
| 7 | (neurological adj3 model*).tw,kf,id. | **4884** |
| 8 | (head adj3 model*).tw,kf,id. | **12580** |
| 9 | (interferen* adj3 stimulat*).tw,kf,id. | **1381** |
| 10 | transcranial electric* stimulat*.tw,kf,id. | **3342** |
| 11 | ((electric* or current*) adj3 stimulat*).tw,kf,id. | **227044** |
| 12 | or/1-11 | **12664613** |
| 13 | TI stimulat*.tw,kf,id. | **194** |
| 14 | (interferen* adj3 therap*).tw,kf,id. | **3883** |
| 15 | (temporal* adj3 interfer*).tw,kf,id. | **1315** |
| 16 | (interferen* adj3 current*).tw,kf,id. | **1803** |
| 17 | temporal* interfere*.tw,kf,id. | **446** |
| 18 | tTIS.tw,kf,id. | **1206** |
| 19 | or/13-18 | **7898** |
| 20 | 12 and 19 | **2739** |
| 21 | remove duplicates from 20 | **1923** |

###### Cochrane’s Library CENTRAL Database Search

Date Run:	12/12/2024 19:32:53

| **ID** | **Search Hits** |  |
| --- | --- | --- |
| **#1** | MeSH descriptor: [Brain] explode all trees | **18840** |
| **#2** | MeSH descriptor: [Models, Neurological] explode all trees | **287** |
| **#3** | MeSH descriptor: [Clinical Study] explode all trees | **45** |
| **#4** | MeSH descriptor: [Electric Stimulation Therapy] explode all trees | **11536** |
| **#5** | (brain*):ti,ab,kw (Word variations have been searched) | **85874** |
| **#6** | ((neurological NEAR/3 model*)): ti, ab,kw (Word variations have been searched) | **344** |
| **#7** | ((head NEAR/3 model*)):ti,ab,kw (Word variations have been searched) | **161** |
| **#8** | (clinical NEAR/3 (stud* or trial* or project*)):ti,ab,kw (Word variations have been searched) | **814577** |
| **#9** | ((interferen* NEAR/3 stimulat*)):ti,ab,kw (Word variations have been searched) | **175** |
| **#10** | (transcranial electric* stimulat*):ti,ab,kw (Word variations have been searched) | **2275** |
| **#11** | ((electric* or current*) NEAR/3 stimulat*):ti,ab,kw (Word variations have been searched) | **22759** |
| **#12** | #1 OR #2 OR #3 OR #4 OR #5 OR #6 OR #7 OR #8 OR #9 OR #10 OR #11 | **879751** |
| **#13** | (TI stimulat*):ti,ab,kw (Word variations have been searched) | **225** |
| **#14** | ((interferen* NEAR/3 therap*)):ti,ab,kw (Word variations have been searched) | **474** |
| **#15** | ((temporal* NEAR/3 interfer*)):ti,ab,kw (Word variations have been searched) | **39** |
| **#16** | ((interferen* NEAR/3 current*)):ti,ab,kw (Word variations have been searched) | **383** |
| **#17** | (temporal* interfere*):ti,ab,kw (Word variations have been searched) | **262** |
| **#18** | (tTIS):ti,ab,kw (Word variations have been searched) | **27** |
| **#19** | #13 OR #14 OR #15 OR #16 OR #17 OR #18 | **1188** |
| **#20** | #12 AND #19 | **793** |

###### ClinicalTrials.gov, WHO ICTRP, and PubMed Search

**Clinical Trials.gov**

Date Run: 12 December 2024 Number of searches: 1

**Search Terms	Results**

(temporal interference OR temporal interference stimulation OR interferential current therapy OR Interferential current stimulation OR temporal interferential stimulation OR temporal interference electrical stimulation OR interferential electrical stimulation)

**113**

**WHO ICTRP**

Date Run: 12 December 2024 Number of searches: 1

**Search Terms	Results**

(temporal interference OR temporal interference stimulation OR interferential current therapy OR Interferential current stimulation OR temporal interferential stimulation OR temporal interference electrical stimulation OR interferential electrical stimulation)

**124**

**PubMed (Supplementary Search)** Date Run: 12 December 2024 Number of searches: 1

**Search Terms	Results**

("temporal interference"[Title/Abstract] OR "temporal interference	**44**

stimulation"[Title/Abstract] OR "TI stimulation"[Title/Abstract] OR "interference stimulation"[Title/Abstract]) AND ("transcranial stimulation"[Title/Abstract] OR "brain stimulation"[Title/Abstract]) AND (human[MeSH Terms] OR human[Title/Abstract] OR participants[Title/Abstract] OR subjects[Title/Abstract] OR patients[Title/Abstract])

##### Eligibility Criteria

###### Inclusion criteria

Original published peer-reviewed journal articles, editorials, notes, letters, errata, case reports, case series, guidelines, protocols, trial registry records;

If a trial registry record has an associated publication, it is reviewed as a published study and NOT as a trial registry record (i.e., removed from the total count for trial registries);

Studies that administered transcranial temporal interference stimulation (tTIS) in human participants;

Temporal interference stimulation should have been delivered to the scalp/head (i.e., should be transcranial).

Transorbital or peripheral temporal interference stimulation is excluded;

Studies with both healthy participants and participants with any disorder are included;

No age restrictions for the study participants;

tTIS in combination with other treatments (e.g., tTIS + medications; tTIS + psychotherapy) allowed but should be clearly noted;

English language

###### Exclusion criteria

Books, chapters, conference proceedings, abstracts, preprints, reviews, meta-analyses, theses/dissertations;

Transorbital tTIS, peripheral tTIS, cranial electrotherapy stimulation (CES), tDCS, tRNS, and tACS excluded;

Non-English language;

Studies on animal models;

Computational/simulation studies on head models with no human participants;

Biophysics studies;

Electrophysiological, cortical potential, and computational studies measuring the effect of tTIS on the function of neurons (without a clear report of human patient outcomes);

##### List of Extracted Variables

**Bibliographic information and meta-data**

Publication/registration year

Title

Authors

Sponsors

Collaborators

Country

**Participant characteristics**

Age

Sex

Population (healthy or clinical)

Diagnoses (if applicable)

**Study design**

Sample size

Study arms

Intervention model

Allocation

Masking

**Study outcomes**

Clinical or psychometric

MRI

Electrophysiological

Laboratory tests

Biological rhythms

Molecular imaging

Behavioural

Safety

**tTIS parameters**

Carrier frequencies

Envelope frequency

Amplitude

Ramp time

Electrode placement

Stimulation target

Duration of one stimulation session per condition (active or control)

Total number of stimulation sessions completed by one participant (active or control)

Manufacturer of the tTIS device

**Results**

Clinical and psychometric

MRI

Electrophysiological

Laboratory tests

Biological rhythm

Molecular imaging

Behavioural

Safety

##### Quality Assessment

###### eTable S1. Quality Assessment of Controlled Intervention Studies

|  | **1** | **2** | **3** | **4** | **5** | **6** | **7** | **8** | **9** | **10** | **11** | **12** | **13** | **14** | **Total** |
| --- | --- | --- | --- | --- | --- | --- | --- | --- | --- | --- | --- | --- | --- | --- | --- |
| Ma et al. (2022) | Y | Y | CD | N | N | Y | Y | Y | Y | Y | Y | Y | Y | Y | **11** |
| Piao et al. (2022) | Y | CD | CD | N | N | Y | Y | Y | Y | Y | Y | Y | Y | N | **9** |
| von Conta et al. (2022) | Y | CD | CD | CD | CD | Y | Y | Y | Y | Y | Y | Y | Y | Y | **10** |
| Zhang et al. (2022) | Y | Y | Y | N | N | Y | Y | Y | Y | Y | Y | N | Y | Y | **11** |
| Zhu et al. (2022) | Y | CD | CD | Y | CD | Y | Y | Y | Y | Y | Y | N | Y | Y | **10** |
| Iszak et al. (2023) | Y | N | CD | CD | CD | Y | Y | Y | Y | Y | Y | N | Y | Y | **9** |
| Violante et al. (2023) | Y | Y | CD | N | N | Y | Y | Y | Y | Y | Y | Y | Y | N | **9** |
| Wessel et al. (2023) | Y | Y | CD | Y | CD | Y | Y | Y | Y | Y | Y | Y | Y | N | **11** |
| Beanato et al. (2024) | Y | Y | CD | Y | CD | Y | Y | Y | Y | Y | Y | N | Y | Y | **11** |
| Demchenko et al. (2024) | Y | Y | Y | Y | CD | Y | Y | Y | Y | Y | Y | N | Y | N | **11** |
| Modak et al. (2024) | Y | CD | CD | N | N | Y | Y | Y | Y | Y | Y | N | Y | N | **8** |
| Thiele et al. (2024) | Y | Y | Y | Y | Y | NA | NA | NA | NA | Y | Y | Y | Y | NA | **9** |
| Vassiliadis et al. (2024a)^1^ | Y | Y | CD | CD | CD | Y | Y | Y | Y | Y | Y | N | Y | Y | **10** |
| Vassiliadis et al. (2024b)^2^ | Y | CD | CD | CD | CD | Y | Y | Y | Y | Y | Y | Y | Y | N | **9** |
| Wang et al. (2024) | Y | CD | CD | Y | CD | N | Y | Y | Y | Y | Y | N | Y | Y | **9** |
| Yang et al. (2024a)^3^ | Y | Y | CD | Y | CD | Y | Y | Y | Y | Y | Y | Y | Y | N | **11** |
| Zheng et al. (2024) | Y | Y | Y | Y | Y | Y | Y | Y | Y | Y | Y | N | Y | N | **12** |
| Zhu et al. (2024) | Y | Y | Y | Y | Y | Y | Y | Y | Y | Y | Y | Y | Y | Y | **14** |
| **Q1**: Was the study described as randomized, a randomized trial, a randomized clinical trial, or an RCT?  **Q2**: Was the method of randomization adequate (i.e., use of randomly generated assignment?)  **Q3**: Was the treatment allocation concealed (so that assignments could not be predicted)?  **Q4**: Were study participants and providers blinded to treatment group assignment?  **Q5**: Were the people assessing the outcomes blinded to the participants’ group assignments?  **Q6**: Were the groups similar at baseline on important characteristics that could affect outcomes (e.g., demographics, risk factors, co-morbid conditions)?  **Q7**: Was the overall drop-out rate from the study at endpoint 20% or lower of the number allocated to treatment?  **Q8**: Was the differential drop-out rate (between treatment groups) at endpoint 15 percentage points or lower?  **Q9**: Was there high adherence to the intervention protocols for each treatment group?  **Q10**: Were other interventions avoided or similar in the groups (e.g., similar background treatments)?  **Q11**: Were outcomes assessed using valid and reliable measures, implemented consistently across all study participants?  **Q12**: Did the authors report that the sample size was sufficiently large to be able to detect a different in the main outcome between groups with at least 80% power?  **Q13**: Were outcomes reported or subgroups analyzed prespecified (i.e., identified before analyses were conducted)?  **Q14**: Were all randomized participants analyzed in the group to which they were originally assigned (i.e., did they use an intention-to-treat analysis)?  Y, yes; N, no; CD, cannot determine; NA, not applicable  Total score is calculated as a sum of Y responses out of 14  ^1^ Vassiliadis et al. (2024). *J Neural Eng*.  ^2^ Vassiliadis et al. (2024). *Nat Hum Behav*.  ^3^ Yang et al. (2024). *Mov Disord*. | | | | | | | | | | | | | | | |

###### eTable S2. Quality Assessment of Before-After (Pre-Post) Studies With No Control Group

|  | **1** | **2** | **3** | **4** | **5** | **6** | **7** | **8** | **9** | **10** | **11** | **12** | **Total** |
| --- | --- | --- | --- | --- | --- | --- | --- | --- | --- | --- | --- | --- | --- |
| Liu et al. (2024) | Y | Y | Y | CD | N | Y | Y | Y | Y | N | N | NA | **7** |
| Yang et al. (2024b)^1^ | Y | Y | Y | CD | N | Y | Y | N | Y | N | Y | NA | **7** |
| **Q1**: Were the study question and objective clearly stated?  **Q2**: Were eligibility/selection criteria for the study population prespecified and clearly described?  **Q3**: Were the participants in the study representative of those who would be eligible for the test/service/intervention in the general or clinical population of interest?  **Q4**: Were all eligible participants that met the prespecified entry criteria enrolled?  **Q5**: Was the sample size sufficiently large to provide confidence in the findings?  **Q6**: Was the test/service/intervention clearly described and delivered consistently across the study population?  **Q7**: Were the outcome measures prespecified, clearly defined, valid, reliable, and assessed consistently across all study participants?  **Q8**: Were the people assessing the outcomes blinded to the participants’ exposures/interventions?  **Q9**: Was the loss to follow-up after baseline 20% or less? Were those lost to follow-up accounted for in the analysis?  **Q10**: Did the statistical methods examine changes in outcome measures from before to after the intervention? Were statistical tests done that provided p values for the pre-to-post changes?  **Q11**: Were outcome measures of interest taken multiple times before the intervention and multiple times after the intervention (i.e., did they use an interrupted time-series design)?  **Q12**: If the intervention was conducted at a group level (e.g., a whole hospital, a community, etc.), did the statistical analysis take into account the use of individual-level data to determine effects at the group level?  Y, yes; N, no; CD, cannot determine; NA, not applicable  Total score is calculated as a sum of Y responses out of 11, since Q12 was rated as N/A for both studies  **^1^** Yang et al. (2024). *Brain Stimul*. | | | | | | | | | | | | | |

Reference: National Heart, Lung, and Blood Institute (2019). Study Quality Assessment Tools

<https://www.nhlbi.nih.gov/health-topics/study-quality-assessment-tools>

###### eTable S3. Quality of the Evidence

| **Study** | **Quality Rating** |
| --- | --- |
| Ma et al. (2022) | 1 |
| Piao et al. (2022) | 1 |
| von Conta et al. (2022) | 1 |
| Zhang et al. (2022) | 1 |
| Zhu et al. (2022) | 1 |
| Iszak et al. (2023) | 4 |
| Violante et al. (2023) | 1 |
| Wessel et al. (2023) | 1 |
| Beanato et al. (2024) | 1 |
| Demchenko et al. (2024) | 1 (Study protocol) |
| Liu et al. (2024) | 4 |
| Modak et al. (2024) | 2 |
| Thiele et al. (2024) | 1 |
| Vassiliadis et al. (2024a)^a^ | 1 |
| Vassiliadis et al. (2024b)^b^ | 1 |
| Wang et al. (2024) | 1 |
| Yang et al. (2024a)^c^ | 1 |
| Yang et al. (2024b)^d^ | 4 |
| Zheng et al. (2024) | 1 |
| Zhu et al. (2024) | 1 |
| Rating scheme adapted from the Oxford Centre for Evidence-based Medicine  1 = Properly powered and conducted randomized controlled trial; systematic review with meta-analysis  2 = Well-designed controlled trial without randomization; prospective comparative cohort trial  3 = Case-control studies; retrospective cohort study  4 = Case series with or without intervention; cross-sectional study  5 = Opinion of respected authorities; case reports  ^a^ Vassiliadis et al. (2024). *J Neural Eng*.  ^b^ Vassiliadis et al. (2024). *Nat Hum Behav*.  ^c^ Yang et al. (2024). *Mov Disord*.  ^d^ Yang et al. (2024). *Brain Stimul*. | |

### eResults

###### Research Design

Of the 20 published studies, 18 (90%) were randomized controlled trials (RCTs),1–18 among which 11 (55%) used a within-subjects crossover design (**eTable S4** and **Figure 2B** in the **Manuscript**).1,3,5–9,12,15–17 The predominant theme of the 20 published studies was a mechanistic exploration of transcranial temporal interference stimulation (tTIS) effects, featured in 18 studies (90%),1–10,12–16,18–20 whereas tTIS as a potential treatment was explored in 4 studies (20%).11,17,19,20 Of the 28 clinical trials, 15 (54%) are currently active (recruiting or not yet recruiting). Similar to the published studies, the predominant theme of the ongoing clinical trials is a mechanistic exploration of tTIS effects (*n* = 16; 57%). However, more clinical trials are shifting towards investigating tTIS as a potential treatment for neurological or psychiatric disorders (*n* = 17; 61%), and 1 trial (4%) is exploring device feasibility.

Sixteen published studies administered tTIS as part of a single-session protocol,1–7,9,10,12,13,15–17,19,20 whereas 4 utilized multi-session protocols ranging from 2 to 10 sessions (**eTable S5**).8,11,14,18 Among multi-arm studies (*n* = 19; 90%), four (20%) compared tTIS across different envelope frequencies,1,2,10,15 seven (35%) used transcranial alternating current stimulation (tACS) as an active control,3,4,6,8,13,14,16 two (10%) used transcranial direction current stimulation (tDCS) as an active control,^5,9^ and 14 (70%) used 0 mA current stimulation as a sham control.1–4,6,7,10– 15,17,18 Twelve published tTIS studies (57%) featured crossover assignments to allocate conditions,1,3,5–9,12,15–17,19 with Liu et al. (2024)^19^ having both single-group and crossover assignments. Thirteen studies (65%) were double-blind.3,5,8,9,11–19 Among the ongoing trials, 16 trials (57%) feature a multi-session tTIS protocol ranging from 2-40 sessions. For the trials with multiple arms (*n* = 21; 75%), six (21%) feature different tTIS envelope frequency or target comparisons, four (14%) use tACS as an active control, one (4%) features tDCS as an active control, and 18 (64%) feature 0 mA stimulation as a sham control. Half of the upcoming tTIS trials (*n* = 14; 50%) are double-blind.

###### Participant Characteristics

Of the 20 published tTIS studies, 5 involved clinical populations (25%): 3 enrolled participants with Parkinson’s disease (PD) or essential tremor (ET),^17,19,20^ 1 study focused on the assessment of safety among healthy individuals and patients with traumatic brain injury (TBI),^14^ and 1 published protocol focuses on applying tTIS in individuals with major depressive disorder (MDD).^11^ In the PD studies, participants exhibited mild-to-moderate disability. A double-blind randomized controlled trial (RCT) by Yang et al. (2024a)^17^ reported baseline Movement Disorder Society–Unified Parkinson’s Disease Rating Scale, Part III (MDS-UPDRS-III) and Hoehn & Yahr scores of

41.8 (2.4) and 2.0 (0.4), respectively, while an open-label trial by Yang et al. (2024b)^20^ reported baseline scores of 39 (3.8) and 2.4 (0.4), respectively. Across these studies, participants had a mean illness duration of 8.9 years.^17,20^ Notably, none of the published studies involving clinical populations included a healthy control group.

In contrast, ongoing tTIS clinical trials more frequently enroll clinical populations: of 28 ongoing trials, 9 include healthy participants (32%) (**eTable S4**). The use of tTIS for MDD has expanded, with 6 trials enrolling individuals with MDD (21%) and 1 trial investigating subthreshold MDD (4%). Additional ongoing trials enroll participants with epilepsy (*n* = 3; 11%), PD (*n* = 2; 7%), substance use or gambling disorder (*n* = 2; 7%), and 1 trial each (4%) enrolling individuals with bipolar disorder, cerebral palsy, post-stroke cognitive impairment, Alzheimer’s disease, and prolonged disorders of consciousness.

Published studies and ongoing clinical trials show comparable participant age and sex distributions. Most published studies focused on adults: 14 of 20 studies (70%) included only adult participants (18–64 years old),1–7,9,10,12,13,15,16,18 2 studies (10%) included only older adults (≥65 years old),^17,20^ and 4 studies (20%) included both adults and older adults.8,11,14,19 Across studies reporting age data, the weighted mean age of participants who received either active tTIS or control was 28.6 years.1,2,4,5,7–10,12–20 Sex distribution in published studies was balanced. Eighteen of 20 studies (90%) included both male and female participants (all except Zheng et al. [2024]^18^ and Liu et al. [2024]^19^), with a weighted mean female representation of approximately 46%.1,2,4,5,7–10,12–20 Similarly, most ongoing clinical trials include adult participants, although children (<18 years old) and older adults are more frequently represented. Eleven of 28 trials (39%) enroll only adults, 3 trials (11%) enroll only children, 11 trials (39%) include both adults and older adults, and 3 trials (11%) include both children and adults. All 28 trials (100%) include both male and female participants.

###### Outcomes

###### Safety and Tolerability Outcomes

Sixteen of 20 published tTIS studies (80%) reported on safety or tolerability outcomes, including adverse event (AE) frequency and severity, dropout rates, and sensation ratings.1–4,7,10–20 Among the 10 studies1,2,4,7,10,14,15,17,19,20 that differentiated AEs between tTIS and control groups, tingling (*P* < 0.05) and itching (*P* < 0.001) were reported significantly more frequently with tTIS (**Table 2** in the **Manuscript**). These findings align with the only two studies in the review that found statistically significant differences in AEs between tTIS and sham: Violante et al. (2023)^7^ reported increased itchiness at the stimulation site with tTIS (*P* = .02), and Wang et al. (2024)^10^ found that more participants receiving 70 Hz tTIS reported tingling compared to 20 Hz tTIS or sham (*P* = .03). No serious AEs were reported across any study. Electroencephalography (EEG) data showed no epileptic activity (as assessed using automated epilepsy detection software, Encevis), and no electrodes exceeded body temperature. Only one study reported participant withdrawal: a single individual with TBI discontinued due to strong sensations during tTIS.^14^

Three studies (15%)^7,12,14^ evaluated tTIS safety and tolerability using sensation ratings. None found significant differences between tTIS and control. Vassiliadis et al. (2024b)^14^ used a 0-3 scale (none to strong) and found average ratings of 0.98 in healthy participants and 0.83 in those with TBI following 2 mA tTIS, with no significant differences between groups. However, among healthy participants, older adults reported significantly lower sensation ratings than younger adults (*P* = .002). Modak et al. (2024)^12^ used alternating tTIS and sham blocks and found no significant changes in discomfort ratings either between conditions or relative to baseline. Violante et al. (2023)^7^ rated AEs on a 1-4 scale (absent to severe), with no AEs rated as severe (4) and no average intensity exceeding 2 in either condition. They also assessed perception thresholds, finding that sensations were first reported at 1.73-2.25 mA during tTIS, compared to 0.33-0.44 mA for tACS control.

###### Clinical and Psychometric Outcomes

To evaluate the effects of tTIS, we categorized each study based on comparisons between active tTIS and control conditions. Following definitions adapted from Heim et al. (2023),^21^ we applied this framework specifically to clinical, psychometric, and behavioural outcomes. Studies were classified as “Superior to Control” when tTIS demonstrated a statistically significant (*P* < .05) effect on the primary outcome in a direction consistent with the study’s hypothesis compared to a control group. “No Difference” was assigned when no statistically significant differences were observed between tTIS and control conditions or when effects were not in the predicted direction. In cases where studies reported mixed results across multiple outcome measures, we considered the overall direction of effect relative to the *a priori* hypothesis. As none of the early-phase, primary mechanistic studies showed effects opposite to the predicted direction, an “Inferior to Control” category was not included.

*Neurological Symptoms*

To date, peer-reviewed, controlled studies on the clinical efficacy of tTIS have focused exclusively on patients with PD and ET. Among these, only Yang et al. (2024a)^17^ employed a randomized, double-blind, sham- controlled design. A case series by Liu et al. (2024)^19^ involving 2 PD and 1 ET patients included tACS as an active control in one case. The clinical efficacy of tTIS in 2 studies^17,19^ is summarized in **eTable S9**, while Yang et al. (2024b)^20^ is summarized separately due to its uncontrolled, within-subjects design (**eTable S10**).

All three studies assessed motor symptoms using the MDS-UPDRS-III.^22^ A double-blind RCT by Yang et al. (2024)^17^ targeted the right globus pallidus internus (GPi) in 12 mild PD patients (Hoehn & Yahr: 1.5-2.5) during a medication-ON state (≥1 hour post-medication). tTIS was delivered at a 130 Hz envelope frequency, resulting in significant improvements in total MDS-UPDRS-III scores (-6.64 points, 14.7%, *P* = .02) compared to sham. Notably, there were significant reductions in bradykinesia (23.5%, *P* = .01) and tremor (15.3%, *P* = .01). Improvements were greater on the left side (contralateral to the right GPi) for both overall scores (*P* = .05) and bradykinesia (*P* < .01).

Liu et al. (2024)^19^ applied 130 Hz tTIS to the bilateral substantial nigra (SN) in two participants with PD (tremor-dominant) and one with ET. In the single PD participant who completed both active and tACS control sessions, resting tremor amplitude in the right upper limb improved from 1-3 cm at baseline to <1 cm in the active tTIS condition, with no change during the control condition. No statistical analyses were reported due to the small sample size.

In an open-label trial by Yang et al. (2024b),^20^ tTIS was delivered to the unilateral subthalamic nucleus (STN) contralateral to the more affected side in 8 patients with mild-to-moderate PD (Hoehn & Yahr 2-3) in a medication- OFF state (≥12 hours withdrawn). As in other studies, a 130 Hz envelope frequency was used. Unlike the group-based approach in a double-blind RCT by Yang et al. (2024a),^17^ this study employed magnetic resonance imaging (MRI)- guided individualized montages and computational modelling to optimize targeting. tTIS yielded a mean reduction of 11.1 points (27.5%) in the MDS-UPDRS-III immediately post-stimulation, with sustained effects at 60 minutes (10.1 points, 25.0%). Although statistical tests were not reported, our analysis revealed medium-to-large Hedge’s *g* effect sizes (-0.73 to -0.92) for total scores (**eTable S10**). The most robust effects were observed for bradykinesia (Hedge’s *g* range: -0.72 to -0.76) and rigidity (Hedge’s *g* range: -0.66 to -0.88), with smaller effects for tremor (Hedge’s *g* range: -0.17 to -0.45) and axial symptoms (Hedge’s *g* range: -0.28 to -0.39). In terms of lateralization, effect sizes were larger for ipsilateral changes relative to the stimulated STN (e.g., Hedge’s *g* = -1.06 immediately post-stimulation), but contralateral improvements were also observed (e.g., Hedge’s *g* = -0.44), suggesting bilateral engagement of motor networks.

*Standardized Cognitive and Mood Assessments*

Three studies reported cognitive and mood scale outcomes in healthy participants.^2,10,12^ The Montreal Cognitive Assessment (MoCa) and a five-point Self-Assessment Scale (SAS) were used by Piao et al. (2022)2 to assess the safety of low-current (1 mA, zero-to-peak) tTIS targeting the left primary motor cortex (M1). This protocol was later replicated by Wang et al. (2024)^10^ using high-current (15 mA, zero-to-peak) tTIS for the same target. Modak et al. (2024)^12^ also assessed mood using the Positive and Negative Affect Schedule (PANAS) as a secondary outcome following 20 Hz tTIS of the left caudate nucleus.

Piao et al. (2022)^2^ found no significant differences between tTIS and sham groups on the Visual Analog Mood Scale (VAMS-R) or SAS for the low-current tTIS. In contrast, Wang et al. (2024)^10^ reported a significant reduction in Calmness ratings on the SAS for participants who received high-current 20 Hz tTIS compared to those who received 70 Hz or sham stimulation (*P* = .04). While Modak et al. (2024)^12^ did not report between-group comparisons for PANAS, a significant reduction in negative affect was observed post-stimulation relative to baseline (*P* = .006).

###### Behavioural Outcomes

Thirteen of 20 published studies examined behavioural outcomes.1–4,7,8,10–13,15,16,18 grouped into three primary domains: motor, memory and cognitive performance, and visual (**eTable S11**).

*Motor Outcomes*

Motor function was the most frequently studied domain, with 6 of 13 studies examining motor performance in healthy participants.1,2,8,10,15,18 Four of these targeted the M1 with tTIS.1,2,10,18 Wang et al. (2024)^10^ found that 20 Hz tTIS applied to M1 significantly slowed relation times (RT) in a one-increment task compared to 70 Hz tTIS and sham (group x time, *P* = .046). While neither 20 Hz nor 70 Hz tTIS improved performance on a simple reaction time (SRT) task, a trend toward improvement was noted for 70 Hz (*P* = 0.059). Piao et al. (2022),^2^ using the same stimulation conditions and M1 target, found no significant effects on manual dexterity (Purdue Pegboard Test [PPT]) from pre- to post-stimulation for all conditions. Additionally, they reported no differences in an abbreviated version of the California Computerized Assessment Package (A-CalCAP), which tests psychomotor performance with the SRT, choice reaction time for single digits (CRT), serial pattern matching 1 (SPM1), and serial pattern matching 2 (SPM2) tasks. Ma et al. (2022)^1^ reported that 70 Hz tTIS produced the lowest mean RT in a random reaction time task (RRTT) compared to both sham and 20 Hz tTIS, reaching significance only against sham (*P* = 0.02). They also observed improved first implicit motor learning (FIL) in a serial reaction time task (SRTT) following 20 Hz tTIS (*P* = 0.04). In a 5-day RCT, Zheng et al. (2024)^18^ applied repetitive 20 Hz tTIS to bilateral M1 leg area and reported enhanced vertical jump performance, including countermovement jump height (*P* = .004), squat jump height (*P* = .01), and average continuous jump height (*P* = .004), with no change in postural stability (Y-balance test).

Two studies targeted the striatum and assessed motor learning.^8,15^ Wessel et al. (2023)^8^ used intermittent theta-burst stimulation (iTBS)-patterned tTIS at 100 Hz, reporting improved performance in a 9-digit sequential finger tapping task (SFTT) compared to tACS control (*P* = .01). Vassiliadis et al. (2024)^15^ found that 80 Hz tTIS, but not 20 Hz or sham, disrupted reinforcement-based motor learning in a force-tracking task (*P* = .04). In total, 4 of the 6 studies reported tTIS to be superior to control, suggesting frequency-dependent effects on motor performance.1,8,15,18

*Memory and Cognitive Performance Outcomes*

Five studies assessed the effect of tTIS on cognitive function, focusing on working memory (WM),^3,4,12^ episodic memory,^7^ and spatial memory.^16^ Modak et al. (2024)^12^ found no significant WM differences on a digit span task between 20 Hz tTIS targeting the left caudate nucleus and sham. Zhang et al. (2022)^4^ reported a slight improvement in 3-back task RTs following 6 Hz tTIS of the frontoparietal cortex compared to tACS control (*P* < .05), although this effect was limited to pre-to-post comparisons and not significant during stimulation. No differences were observed for other n-back conditions or stimulation types. Similarly, von Conta et al. (2022)^3^ reported no difference in performance on a simple visual change detection task in participants who received alpha tTIS of the parieto-occipital cortex compared to tACS or sham.

Two studies targeted hippocampal regions to assess episodic and spatial memory.^7,16^ Beanato et al. (2024)^16^ found that iTBS-patterned 100 Hz tTIS to the right hippocampus improved spatial navigation latency in a virtual task relative to continuous theta-burst stimulation (cTBS) tTIS (*P* < .001) or control (*P* = .04), without affecting accuracy. Violante et al. (2023)^7^ applied 5 Hz tTIS (1:3 mA amplitude ratio between the two channels) during face-name encoding, resulting in higher recall accuracy than sham at immediate (*P* = .007) and delayed (*P* = .006) recall, with a 12% increase in odds of correct recall (Bayesian 95% CI [0.02, 0.21]).

*Visual Outcomes*

Two studies assessed visual processing following tTIS to the parieto-occipital cortex.^6,13^ Thiele et al. (2024) used Shepard’s mental rotation task and found that all stimulation groups (tTIS, tACS, sham) improved in accuracy and RT from pre- to post-stimulation (*P* < .001), but with no between-group differences. Iszak et al. (2023)^6^ tested phosphene perception at high intensities (up to 4 mA) and reported no stimulation-induced phosphenes under tTIS or sham conditions.

###### Neuroimaging Outcomes

Eight studies incorporated MRI into their design.5,7–9,11,12,15,16 Von Conta et al. (2022)^3^ used anatomical MRI to support source-level magnetoencephalography (MEG) analysis. In contrast, Demchenko et al. (2023)^11^ proposed a protocol using multiparameter imaging, including anatomical, functional, diffusion, and perfusion imaging, to investigate target engagement of the subgenual anterior cingulate cortex (sgACC), although results are not yet available. To date, no molecular imaging (e.g., positron emission tomography [PET]) studies using tTIS have been conducted.

Seven studies reported functional MRI (fMRI) findings,5,7–9,12,15,16 supporting the ability of tTIS to modulate activity and functional connectivity (FC) in targeted brain regions and networks. There is consistent evidence of target engagement of the left M1,^5,9^ hippocampus,^7,16^ and striatum.^8,12,15^

In the motor domain, two resting-state fMRI studies^5,9^ applying 20 Hz tTIS to left M1 reported increased activation within the sensorimotor network, similar to effects seen with tDCS. Zhu et al. (2024)^9^ additionally found that tTIS reduced the coefficient of variation in dynamic FC of the precentral and postcentral gyri and increased overall FC strength relative to tDCS, suggesting that 20 Hz tTIS may enhance network stability via resonance with intrinsic M1 dynamics.

In the hippocampal domain, Violante et al. (2023)^7^ applied 5 Hz tTIS to the left anterior hippocampus during a face-name associative memory task. Compared to both sham and 1:1 mA amplitude ratio conditions, 1:3 mA tTIS significantly reduced memory-evoked blood-oxygen-level-dependent (BOLD) response and decreased FC between the hippocampus and the anterior-temporal (AT) network, with regionally specific patterns of FC reduction. These findings suggest that amplitude ratio and envelope frequency can shape hippocampal-cortical interactions. Beanato et al. (2024)^16^ applied 100 Hz iTBS-patterned tTIS over the right hippocampus and found reductions in grid cell-like representations in the entorhinal cortex, a signal inferred from hexadirectional modulation of BOLD response during spatial navigation. This pattern is thought to reflect impaired spatial encoding mechanisms, offering novel evidence for tTIS influence on hippocampal-entorhinal circuits.

Striatal-targeted studies further underscore the behavioural relevance of tTIS-induced neuromodulation. Wessel et al. (2023)^8^ used 100 Hz iTBS-patterned tTIS during an SFTT and found increased task-related activation in sensorimotor regions of the putamen compared to tACS control. This activation shift toward sensorimotor areas over time was paralleled by improved task performance, supporting the idea that tTIS can facilitate the reorganization of subcortical motor circuits, particularly during active engagement. Similarly, Vassiliadis et al. (2024a)^15^ showed that 80 Hz tTIS (but not 20 Hz or sham) modulated FC within the striatal-frontal network during a reinforcement-based motor learning task, indicating frequency-specific modulation of reinforcement learning mechanisms. In contrast, Modak et al. (2024)^12^ applied 20 Hz tTIS to the left caudate during a virtual reality navigation task and observed increased BOLD activation in the mid-orbitofrontal cortex and parahippocampal gyrus. However, the study noted limited spatial specificity of stimulation, highlighting a key challenge in subcortical tTIS targeting.

###### Neurophysiological Outcomes

Five studies examined the effect of tTIS on neurophysiological outcomes among healthy individuals.2,3,6,10,13 Among those studies, three administered tTIS at alpha envelope frequencies,^3,6,13^ and two administered tTIS at beta and gamma envelope frequencies.^2,10^ A MEG study by von Conta et al. (2022)^3^ examined the modulation of neurophysiological activity during a simple visual change detection task with tTIS applied to the parieto-occipital cortex at an individual alpha frequency (IAF). On the other hand, an EEG study by Iszak et al. (2023)^6^ evaluated the ability of a single session of 10 Hz (alpha) tTIS to modulate alpha oscillations in the occipital cortex. In both studies, alpha tTIS failed to modulate posterior alpha oscillations.

In contrast, Thiele et al. (2024)^13^ reported changes in alpha oscillations during a mental rotation task following both tTIS and tACS applied to the parieto-occipital cortex at IAF, focusing on event-related desynchronization (ERD) of alpha activity as well as resting-state alpha power. Both tTIS and tACS enhanced task- evoked alpha ERD compared to sham, whereas resting-state alpha power remained unchanged. Lastly, Piao et al. (2022)^2^ and Wang et al. (2024)^10^ explored the impact of tTIS applied to the left M1 at 20 Hz (beta) or 70 Hz (gamma) and reported no epileptic activity or significant changes in EEG band powers.

###### Biochemical and Biophysiological Outcomes

Piao et al. (2022)^2^ and Wang et al. (2024)^10^ examined changes in blood neuron-specific enolase (NSE) levels following tTIS applied to the left M1 at 20 Hz vs. 70 Hz. Findings were mixed, as NSE levels decreased after tTIS in Piao et al. (2022) but remained unchanged in Wang et al. (2024). Among ongoing clinical trials, 3/28 (11%) studies from China plan to collect biochemical and biophysiological outcomes (**eTable S6**). These include changes in gastric electrical indices, heart rate variability, and eye movement indicators with tTIS of the left dorsolateral prefrontal cortex vs. left hippocampus for Alzheimer’s disease (ChiCTR2400090199). Additional planned outcomes include blood biomarkers with tTIS of the right amygdala for MDD (NCT06461260) and factors carried by peripheral blood and exosomes (i.e., brain-derived neurotrophic factor, reelin, hypocretin, and N-Methyl-D-aspartic acid receptor) in a large 400-subject trial for treatment-resistant MDD that will identify relevant tTIS targets using data-driven tools (NCT05777876).

###### eTable S4. Study Design Characteristics of Human tTIS Studies and Ongoing Clinical Trials

| **Study** | **Status** | **Country** | **Study design** | ***n*,**  **total** | ***n*, analysis** | **Population** | **# of study groups** | **Groups** | **Alloc.** | **Assign.** | **Masking** | **Outcomes** |
| --- | --- | --- | --- | --- | --- | --- | --- | --- | --- | --- | --- | --- |
| **Published studies** | | | | | | | | | | | | |
| Ma et al. (2022) | Published | China | WSD, RCT | RRTT: 27; SRTT: 33 | RRTT: 21; SRTT: 29 | Healthy | 3 | tTIS (20 Hz) vs. tTIS (70 Hz) vs. sham | R | Crossover | Single-blind | Safety; RRTT and SRTT performance; target excitability |
| Piao et al. (2022) | Published | China | RCT | 40 | 38 | Healthy | 3 | tTIS (20 Hz) vs. tTIS (70 Hz) vs. sham | R | Parallel | Single-blind | Safety; EEG; laboratory tests; neuropsychological (MoCA, A-CalCAP, PPT, VAMS-R, SAS) |
| von Conta et al. (2022) | Published | Germany | WSD, RCT | 34 | 33 | Healthy | 3 | tTIS vs. tACS vs. sham | R | Crossover | Double-blind | Safety; MEG; visual change detection task |
| Zhang et al. (2022) | Published | China | RCT | 60 | 54 | Healthy | 4 | tTIS vs. tACS vs. sham | R | Parallel | Single-blind | Safety and blinding efficiency; working memory task performance |
| Zhu et al. (2022) | Published | China | WSD, RCT | 40 | 40 | Healthy | 2 | tTIS vs. tDCS | R | Crossover | Double-blind | fMRI target engagement |
| Iszak et al. (2023) | Published | Germany | WSD, RCT | Alpha power study: 10 | Alpha power study: 10 | Healthy | 4 | tTIS vs. tACS vs. carrier control vs. sham | PsR | Crossover | Single-blind | EEG |
| Violante et al. (2023) | Published | UK | WSD, RCT | fMRI: 22; Behavioural: 21 | fMRI: 20; Behavioural: 21 | Healthy | 3 | tTIS (mA ratio 1:1) vs. tTIS (mA ratio 3:1) vs. sham | PsR | Crossover | Single-blind | fMRI target engagement; working memory task performance; safety and blinding efficiency |
| Wessel et al. (2023) | Published | Switzerland | WSD, RCT | fMRI: 15; Behavioural: 30 (15 young, 15 older) | fMRI 14; Behavioural: 30 | Healthy | 2 | tTIS vs. tACS | PsR | Crossover | Double-blind | fMRI target engagement; motor task performance; age effects; attention and fatigue |
| Beanato et al. (2024) | Published | Switzerland | WSD, RCT | 30 | 28 | Healthy | 3 | tTIS (iTBS) vs. tTIS (cTBS) vs. tACS | PsR | Crossover | Double-blind | fMRI target engagement; safety; spatial navigation task performance in VR environment |
| Demchenko et al. (2024) | Published | Canada | RCT | 30 | N/A | MDD | 2 | tTIS vs. sham | R | Parallel | Double-blind | fMRI, ASL, & DTI target engagement; EEG; clinical (HAM  D-17, QIDS-SR-16, GAD-7, WHO-5, SDS); safety and blinding efficiency |
| Liu et al. (2024) | Published | China | WSD, case series | 3 | 3 | PD, ET | 2 | tTIS vs. tACS | NR | Single Group, Crossover | Double-blind | Safety; clinical (MDS-UPDRS-III) |
| Modak et al. (2024) | Published | USA | WSD, RCT | 16 | 16 | Healthy | 3 | tTIS vs. sham (x begin ON vs begin OFF) | PsR | Crossover | Double-blind | fMRI target engagement; neuropsychological (PANAS); working memory task performance; safety |
| Thiele et al. (2024) | Published | Germany | RCT | 67 | 48 | Healthy | 3 | tTIS vs. tACS vs. sham | R | Parallel | Double-blind | Safety, EEG; mental rotation task performance |
| Vassiliadis et al. (2024a)^a^ | Published | Switzerland | WSD, RCT | 24 | 23 | Healthy | 3 | tTIS (20 Hz) vs. tTIS (80 Hz) vs. sham (x reinforcement ON vs. reinforcement oFF) | R | Crossover | Double-blind | fMRI target engagement; neuropsychological (impulsivity – independent delay-discounting questionnaire); motor learning task performance; safety and blinding efficiency |
| Vassiliadis et al. (2024b)^b^ | Published | Switzerland | RCT | 119 | 118 | Healthy, TBI | 6 | tTIS (iTBS) vs. tTIS (cTBS) vs. tACS vs. sham | R | Parallel | Double-blind | Safety and blinding efficiency |
| Wang et al. (2024) | Published | China | RCT | 90 | 88 | Healthy | 3 | tTIS (20 Hz) vs. tTIS (70 Hz) vs. sham | R | Parallel | Single-blind | Safety; EEG; laboratory tests; motor task performance; neuropsychological (MoCA, VAMS-R, SAS) |
| Yang et al. (2024a)^c^ | Published | China | WSD, RCT | 15 | 12 | PD | 2 | tTIS vs. sham | R | Crossover | Double-blind | Safety and blinding efficiency; clinical (MDS-UPDRS-III) |
| Yang et al. (2024b)^d^ | Published | China | PPD | 8 | 8 | PD | 1 | tTIS only | N/A | Single Group | Open-label | Safety and blinding efficiency; clinical (MDS-UPDRS-III) |
| Zheng et al. (2024) | Published | China | RCT | 46 | 40 | Healthy | 2 | tTIS vs. sham | R | Parallel | Double-blind | Safety and blinding efficiency; lower limb motor function (vertical jump test, Y-balance test) |
| Zhu et al. (2024) | Published | China | WSD, RCT | 40 | 32 | Healthy | 2 | tTIS vs. tDCS | R | Crossover | Double-blind | fMRI target engagement |
| **Registered clinical trials** | | | | | | | | | | | | |
| NCT03747601 (2018) | Not yet recruiting | USA | PPD | 20 | N/A | Healthy | 1 | tTIS only | N/A | Single Group | Open-label | fMRI retinotopy; EEG; neurological (Humphrey visual field test, visual discrimination task) |
| NCT06716866 (2019) | Recruiting | USA, Czechia | PPD | 100 | N/A | Epilepsy | 1 | tTIS only | N/A | Single Group | Open-label | iEEG (interictal biomarkers) |
| NCT04099056 (2019) | Recruiting | USA | PPCGD | 300 | N/A | MDD, Healthy | 2 | tTIS vs. rTMS | NR | Parallel | Open-label | fMRI target engagement; effort-based decision-making task performance |
| NCT04432064 (2020) | Recruiting | USA | RCT | 100 | N/A | Tobacco Use Disorder | 5 | tTIS vs. sham; tTIS vs. tDCS vs. sham | R | Parallel | Single-blind | Clinical (nicotine craving) |
| NCT04761471 (2021) | Completed | Switzerland | WSD, RCT | 70 | N/A | Healthy | 1 | tTIS vs. sham | PsR | Crossover | Single-blind | fMRI target engagement; EEG; cognitive task performance; safety |
| NCT05777876 (2023) | Recruiting | China | RCT | 400 | N/A | MDD | 9 | tTIS vs. sham; tTIS vs. observation; Multiple tTIS Arms (A, B, C); Closed-Loop stimulation; HC experimental vs. sham | R | Parallel | Double-blind | fMRI target engagement; EEG; clinical (HAM-D-24, BSI, HAM-A); neuropsychological (THINC-it); laboratory tests; safety |
| NCT05805215 (2023) | Not yet recruiting | Czechia | WSD, RCT | 30 | N/A | Healthy | 3 | tTIS (hippocampus) vs. tTIS (precuneus) vs. sham | R | Crossover | Double-blind | fMRI target engagement; EEG; working memory task performance |
| ChiCTR2300077258 (2023) | Recruiting | China | WSD, RCT | 20 | N/A | PD | 2 | tTIS vs. sham | R | Crossover | Double-blind | Safety and blinding efficiency; neurological (balance and gait biomechanical indices; EMG index; TUG test; DTC, MoCA); clinical (MDS-UPDRS-III, freezing of gait questionnaire); fatigue |
| DRKS00030841 (2023) | Completed | Germany | WSD, RCT | 40 | N/A | PD | 2 | tTIS vs. sham | R | Crossover | Double-blind | Safety; motor task performance; effort/reward task performance |
| JPRN-UMIN000050056 (2023) | Recruiting | Japan | RCT | 200 | N/A | Healthy | 5 | tTIS (insula) vs. tTIS (S1) vs. tACS (insula) vs. tACS (S1) vs. sham | R | Parallel | Double-blind | fMRI target engagement; sensory and motor function |
| NCT06663969 (2024) | Recruiting | China | PPD | 20 | N/A | DRE | 1 | tTIS only | N/A | Single Group | Open-label | SEEG; clinical (subjective feelings and symptoms) |
| NCT06708143 (2024) | Recruiting | China | PPD | 30 | N/A | DRE | 1 | tTIS only | N/A | Single Group | Open-label | Clinical (seizure count per month, seizure responder rate, QOLIE-31, incidence of SUDEP); safety; neurological (MMSE, MoCA) |
| ChiCTR2400090858 (2024) | Not yet recruiting | China | RCT | 78 | N/A | DoCs | 2 | tTIS vs. sham | R | Parallel | Single-blind | EEG; EMG; fNIRS; anatomical MRI; clinical (CRS-R) |
| ChiCTR2400090777 (2024) | Not yet recruiting | China | RCT | 90 | N/A | MDD | 3 | tTIS (alpha) vs. tTIS (theta) vs. sham | R | Parallel | Double-blind | Clinical (depressive symptoms); reinforcement learning task performance |
| ChiCTR2400085831 (2024) | Not yet recruiting | China | RCT | 250 | N/A | MDD | 3 | tTIS vs. tACS vs. sham | R | Parallel | Double-blind | Clinical (HAM-D-17, HAM-A, PHQ-9, GAD-7, YMRS, HCL-32) |
| ChiCTR2400090107 (2024) | Not yet recruiting | China | RCT | 60 | N/A | Dyskinetic cerebral palsy | 3 | tTIS (putamen) vs. tTIS (GPi) vs. sham | R | Parallel | Double-blind | EEG; MRI; neurological (gross and fine motor function, pediatric balance scale, WeeFIM, gait analysis) |
| ChiCTR2400090199 (2024) | Not yet recruiting | China | WSD | 10 | N/A | AD | 2 | tTIS (left dlPFC) vs. tTIS (left hippocampus) | NR | Crossover | ns | Clinical (ADAS-Cog, CDR, PSQI, GAD-7, PHQ-9); neuropsychological (MMSE, MoCA, eye movement indicators); EEG; PET-MRI target engagement; MRI; biological (HRV, gastric electrical indices) |
| NCT06452849 (2024) | Recruiting | China | RCT | 60 | N/A | MDD | 2 | tTIS (left dlPFC) vs. tTIS (amygdala) | R | Parallel | Double-blind | Clinical (HAM-D-17, HAM-A, MADRS, PHQ-9, GAD-7, SHAPS, PSQI, SF-36, WHOQOL-BREF, ARI, OSI) |
| NCT06461260 (2024) | Recruiting | China | PPD | 30 | N/A | MDD | 1 | tTIS only | N/A | Single Group | Open-label | Clinical (HAM-D-17, HAM-A, SHAPS, SF-36, WHOQOL-BREF, PSQI, GAD-7, QIDS-SR-16, MADRS); laboratory tests; neuropsychological (THINC-it) |
| NCT06467422 (2024) | Not yet recruiting | China | RCT | 30 | N/A | Gambling addiction | 2 | tTIS vs. tACS | R | Parallel | Double-blind | fMRI target engagement; clinical (gambling craving change, impulsivity); sequential decision-making task performance |
| NCT06477276 (2024) | Recruiting | China | RCT | 92 | N/A | MDD | 2 | tTIS vs. sham | R | Parallel | Double-blind | Clinical (HAM-D-17, HAM-D-6, SHAPS, QIDS-SR-16, HAM-A, PSQI, SF-36, WHOQOL-BREF) |
| NCT06516991 (2024) | Recruiting | China | PPD | 20 | N/A | Bipolar disorder | 1 | tTIS only | N/A | Single Group | Open-label | Clinical (HAM-D-17, MADRS, HAM-A, YMRS, SHAPS, TEPS) |
| NCT06547086 (2024) | Recruiting | USA | WSD, RCT | Phase I: 55; Phase II: 24 | N/A | Healthy | 2 | tTIS vs. no stimulation | R | Crossover | Single-blind | Neuropsychological (nap duration, REM sleep, SSS, REST-Q), neurophysiological (sleep spindles, sawtooth wave frequency); vigilance task score performance; emotion regulation task performance |
| NCT06601686 (2024) – Phase I | Recruiting | USA | RCT | 12 | N/A | Healthy | 3 | tTIS vs. tES vs. sham | R | Parallel | Single-blind | EEG; fMRI target engagement; neuropsychological and health (Ego-Disengagement Scale, BFAS, FLS, PROMIS Anxiety & Depression, PSS, QEWB, PWB, IQSS Flourishing Measure, MPoD, EQ, SRIS, NAS, NTS, FFMQ, DDS, RQ, ULS, SCS, ASQ, EIS, BIS-11, DII, VVIQ); implicit association task performance; rapid discrimination task performance; spontaneous mentation task performance; automatic emotion regulation task performance |
| NCT06601699 (2024) – Phase II | Not yet recruiting | USA | RCT | 48 | N/A | Healthy | 3 | tTIS vs. tES vs. sham | R | Parallel | Single-blind | EEG; neuropsychological and health (Ego-Disengagement Scale, BFAS, FLS, PROMIS Anxiety & Depression, PSS, QEWB) |
| NCT06607432 (2024) | Not yet recruiting | USA | WSD, RCT | 30 | N/A | Healthy | 4 | tTIS (0.3 V/m) vs. tTIS (0.35 V/m) vs. tTIS (0.4 V/m) vs. sham | R | Crossover | Double-blind | fMRI target engagement; safety; EEG; target tissue levels of N-acetyl aspartate and glutathione |
| NCT06267521 (2024) | Recruiting | USA | PPCGD | 48 | N/A | Healthy | 4 | tTIS only + sham meditation VS. sham + meditation VS. tTIS (low-frequency) + meditation VS. tTIS (high-frequency) + meditation | NR | Parallel | Single-blind | Clinical (PHQ-9, GAD-7, suicide, PCL-5, B-SCS, PASIS); neuropsychological and health (Healthy Minds index, PSS, ESQ, CFI, DERS-18, PROMIS Sleep Disturbance, WHO-5, RSQ, SDQ, PSQI, NIH Toolbox Loneliness, FFMQ, DDS, PROMIS Meaning & Purpose, D-WAI; PTQ; EQ-D; experience sampling/ecological momentary assessment); fMRI target engagement; death implicit association task performance; reversal learning task performance; multi-source interference task performance; emotional Stroop task performance; meteor mission task performance; emotional persistence task performance; change your mind task performance; spectral power density during sleep |
| ChiCTR2400081207 (2024) | Not yet recruiting | China | RCT | 36 | N/A | Post-Stroke Cognitive Impairment | 2 | tTIS vs. tACS | R | Parallel | Single-blind | Neurological (MoCA, N-back task, Shape Trail Test, Digit Span Memory Test); fNIRS |
| **Abbreviations:** A-CalCAP = California Computerized Assessment Package, Abbreviated version; AD = Alzheimer’s disease; ADAS-Cog = Alzheimer’s Disease Assessment Scale – Cognitive subscale; Alloc. = Allocation; ARI = Apathy Rating Inventory; ASL = Arterial spin labeling; ASQ = Attachment Style Questionnaire; Assign. = Assignment; BFAS = Big Five Aspects Scale; BIS-11 = Barratt Impulsiveness Scale; B-SCS = Brief Suicide Cognitions Scale; BSI = Beck Scale of Suicidal Ideation; CDR = Clinical Dementia Rating Scale; CFI = Cognitive Flexibility Inventory; CRS-R = Coma Recovery Scale, Revised; cTBS = Continuous theta-burst stimulation; DDS = Drexel Defusion Scale; DERS-18 = Difficulties in Emotion Regulation Scale-18; DII = Dickman Impulsivity Inventory; dlPFC = Dorsolateral prefrontal cortex; DoCs = Disorders of consciousness; DRE = Drug-resistant epilepsy; DTC = Dual-task cost; DTI = Diffusion tensor imaging; D-WAI = Digital Working Alliance Inventory; EEG = Electroencephalography; EIS = Emotional Intelligence Scale; EMG = Electromyography; EQ = Experiences Questionnaire; ESQ = Emotional Styles Questionnaire; ET = Essential tremor; FFMQ = Five Facet Mindfulness Questionnaire; FLS = Fulfilled Life Scale; fMRI = Functional magnetic resonance imaging; fNIRS = Functional near-infrared spectroscopy; GAD-7 = Generalized Anxiety Disorder 7-item; GPi = Globus pallidus internus; HAM-A = Hamilton Anxiety Rating Scale; HAM-D-16 = Hamilton Depression Rating Scale 6-Item; HAM-D-17 = Hamilton Depression Rating Scale 17-Item; HAM-D-24 = Hamilton Depression Rating Scale 24-Item; HC = Healthy controls; HCL-32 = Hypomania Checklist-32; HRV = Heart rate variability; iEEG = Intracranial electroencephalography; iTBS = Intermittent theta-burst stimulation; IQSS = Institute for Qualitative Social Science; MADRS = Montgomery-Åsberg Depression Rating Scale; MDD = Major depressive disorder; MEG = Magnetoencephalography; MDS-UPDRS-III = Movement Disorder Society-Unified Parkinson’s Disease Rating Scale, Part III; MMSE = Mini-Mental State Examination; MoCA = Montreal Cognitive Assessment; MPoD = Metacognitive Processes of Decentering Scale; MRI = Magnetic resonance imaging; NAS = Non-Attachment Scale; NIH = National Institutes of Health; NTS = Non-Attachment to Self Scale; N/A = Not applicable; n = Sample size; NR = Non-randomized; ns = Not specified; PANAS = Positive and Negative Affect Schedule; PASIS = Passive and Active Suicidal Ideation Scale; PCL-5 = PTSD Checklist for DSM-5; PD = Parkinson’s disease; PET = Positron emission tomography; PHQ-9 = Patient Health Questionnaire-9; PROMIS = Patient-Reported Outcomes Measurement Information System; PSQI = Pittsburgh Sleep Quality Index; PSS = Perceived Stress Scale; PTQ = Perseverative Thinking Questionnaire; PPT = Purdue Pegboard Test; PPCGD = Pretest-posttest control group design; PPD = Pretest-posttest design; PsR = Pseudorandomized; PWB = Psychological Well-Being; QEWB = Questionnaire for Eudaimonic Well-Being; QIDS-SR-16 = Quick Inventory of Depressive Symptomatology 16-Item; QOLIE-31 = Quality of Life in Epilepsy-31 Inventory; R = Randomized; RCT = Randomized controlled trial; REM = Rapid eye movement; REST-Q = Recovery-Stress Questionnaire; RQ = Relationships Questionnaire; RSQ = Restorative Sleep Questionnaire; RRTT = Random reaction time task; rTMS = Repetitive transcranial magnetic stimulation; S1 = Primary somatosensory cortex; SAS = Self-Assessment Scale; SCS = Social Connectedness Scale; SDS = Sheehan Disability Scale; SDQ = Sleep Depth Questionnaire; SEEG = Stereoelectroencephalography; SF-26 = 36-Item Short Form Health Survey; SHAPS = Snaith-Hamilton Pleasure Scale; SRTT = Serial reaction time task; SSS = Stanford Sleepiness Scale; SRIS = Self-Reflection and Insight Scale; SUDEP = Sudden Unexpected Death in Epilepsy; TBI = Traumatic brain injury; tACS = Transcranial alternating current stimulation; tDCS = Transcranial direct current stimulation; TEPS = Temporal Experience of Pleasure Scale; tES = Transcranial electrical stimulation; tTIS = Transcranial temporal interference stimulation; TUG test = Timed Up and Go test; ULS = UCLA Loneliness Scale; USA = United States of America; VAMS-R = Visual Analog Mood Scale, Revised; VR = Virtual reality; VVIQ = Vividness of Visual Imagery Questionnaire; WeeFIM = Functional Independence Measure for Children; WHO-5 = World Health Organization Five Well-being Index; WHOQOL-BREF = World Health Organization Quality of Life – Brief; WSD = Within-subjects design; YMRS = Young Mania Rating Scale.  ^a^ Vassiliadis et al. (2024). *J Neural Eng*.  ^b^ Vassiliadis et al. (2024). *Nat Hum Behav*.  ^c^ Yang et al. (2024). *Mov Disord*.  ^d^ Yang et al. (2024). *Brain Stimul*. | | | | | | | | | | | | |

###### eTable S5. Collected Outcomes in Published Human tTIS Studies

| **Study** | **Clinical and**  **psychometric** | **MRI** | **Electro-**  **physiological** | **Laboratory**  **tests** | **Biological**  **rhythms** | **Molecular**  **imaging** | **Behavioural** | **Safety** |
| --- | --- | --- | --- | --- | --- | --- | --- | --- |
| Ma et al. (2022) | No | No | Yes | No | No | No | Yes | Yes |
| Piao et al. (2022) | Yes | No | Yes | Yes | No | No | Yes | Yes |
| von Conta et al. (2022) | No | No | Yes | No | No | No | Yes | Yes |
| Zhang et al. (2022) | No | No | No | No | No | No | Yes | Yes |
| Zhu et al. (2022) | No | Yes | No | No | No | No | No | No |
| Iszak et al. (2023) | No | No | Yes | No | No | No | No | No |
| Violante et al. (2023) | No | Yes | No | No | No | No | Yes | Yes |
| Wessel et al. (2023) | No | Yes | No | No | Yes | No | Yes | No |
| Beanato et al. (2024) | No | Yes | No | No | No | No | Yes | Yes |
| Demchenko et al. (2024) | Yes | Yes | Yes | No | No | No | Yes | Yes |
| Liu et al. (2024) | Yes | No | No | No | No | No | No | Yes |
| Modak et al. (2024) | Yes | Yes | No | No | No | No | Yes | Yes |
| Thiele et al. (2024) | No | No | Yes | No | No | No | Yes | Yes |
| Vassiliadis et al. (2024a)^a^ | No | Yes | No | No | No | No | Yes | Yes |
| Vassiliadis et al. (2024b)^b^ | No | No | No | No | No | No | No | Yes |
| Wang et al. (2024) | Yes | No | No | Yes | No | No | Yes | Yes |
| Yang et al. (2024a)^c^ | Yes | No | No | No | No | No | No | Yes |
| Yang et al. (2024b)^d^ | Yes | No | No | No | No | No | No | Yes |
| Zheng et al. (2024) | No | No | No | No | Yes | No | Yes | Yes |
| Zhu et al. (2024) | No | Yes | No | No | No | No | No | No |
| **Total “Yes”, n (%)** | **7 (35%)** | **8 (40%)** | **6 (30%)** | **2 (10%)** | **2 (10%)** | **0 (0%)** | **13 (65%)** | **16 (80%)** |

^a^ Vassiliadis et al. (2024). *J Neural Eng*.

^b^ Vassiliadis et al. (2024). *Nat Hum Behav*.

^c^ Yang et al. (2024). *Mov Disord*.

^d^ Yang et al. (2024). *Brain Stimul*.

###### eTable S6. Collected Outcomes in Ongoing tTIS Clinical Trials

| **Study** | **Clinical and**  **psychometric** | **MRI** | **Electro-**  **physiological** | **Laboratory**  **tests** | **Biological**  **rhythms** | **Molecular**  **imaging** | **Behavioural** | **Safety** |
| --- | --- | --- | --- | --- | --- | --- | --- | --- |
| NCT03747601 (2018) | No | Yes | Yes | No | No | No | Yes | No |
| NCT06716866 (2019) | Yes | No | Yes | No | No | No | No | No |
| NCT04099056 (2019) | No | Yes | No | No | No | No | Yes | No |
| NCT04432064 (2020) | Yes | No | No | No | No | No | No | No |
| NCT04761471 (2021) | No | Yes | Yes | No | No | No | Yes | Yes |
| NCT05777876 (2023) | Yes | Yes | Yes | Yes | No | No | Yes | Yes |
| NCT05805215 (2023) | No | Yes | Yes | No | No | No | Yes | No |
| ChiCTR2300077258 (2023) | Yes | No | Yes | No | Yes | No | Yes | Yes |
| DRKS00030841 (2023) | No | No | No | No | No | No | Yes | Yes |
| JPRN-UMIN000050056 (2023) | Yes | No | No | No | No | No | Yes | No |
| NCT06663969 (2024) | Yes | No | Yes | No | No | No | No | No |
| NCT06708143 (2024) | Yes | No | No | No | No | No | Yes | Yes |
| ChiCTR2400090858 (2024) | Yes | Yes | Yes | No | No | No | Yes | No |
| ChiCTR2400090777 (2024) | Yes | No | No | No | No | No | Yes | No |
| ChiCTR2400085831 (2024) | Yes | No | No | No | No | No | No | No |
| ChiCTR2400090107 (2024) | Yes | Yes | Yes | No | No | No | No | No |
| ChiCTR2400090199 (2024) | Yes | Yes | Yes | Yes | Yes | Yes | Yes | No |
| NCT06452849 (2024) | Yes | No | No | No | No | No | No | No |
| NCT06461260 (2024) | Yes | No | No | Yes | No | No | Yes | No |
| NCT06467422 (2024) | Yes | Yes | No | No | No | No | Yes | No |
| NCT06477276 (2024) | Yes | No | No | No | Yes | No | No | No |
| NCT06516991 (2024) | Yes | No | No | No | No | No | No | No |
| NCT06547086 (2024) | No | No | Yes | No | Yes | No | Yes | No |
| NCT06601686 (2024) | Yes | Yes | Yes | No | No | No | Yes | No |
| NCT06601699 (2024) | Yes | No | Yes | No | No | No | Yes | No |
| NCT06607432 (2024) | No | Yes | Yes | No | No | No | No | Yes |
| NCT06267521 (2024) | Yes | Yes | No | No | Yes | No | Yes | No |
| ChiCTR2400081207 (2024) | Yes | No | Yes | No | No | No | Yes | No |
| NCT03747601 (2018) | No | Yes | Yes | No | No | No | Yes | No |
| **Total “Yes”, n (%)** | **21 (75%)** | **12 (43%)** | **15 (54%)** | **3 (11%)** | **5 (18%)** | **1 (4%)** | **19 (68%)** | **6 (21%)** |

###### eTable S7. Session Duration and Number of Sessions per Condition in Human tTIS Studies

| **Study** | **Session duration per condition** | **Number of sessions per participant** |
| --- | --- | --- |
| Ma et al. (2022) | 30 min | 3 (1 20 Hz tTIS; 1 70 Hz tTIS; 1 sham) |
| Piao et al. (2022) | 30 min (3 blocks x 10 min) | 1 |
| von Conta et al. (2022) | 20 min | 3 (1 tTIS; 1 tACS; 1 sham) |
| Zhang et al. (2022) | 15 min | 1 |
| Zhu et al. (2022) | 20 min | 2 (1 tTIS; 1 tDCS) |
| Iszak et al. (2023) | 10 min (5 min eyes open + 5 min eyes closed) | 2 (1 tTIS + control; 1 tACS + control) |
| Violante et al. (2023) | fMRI: 1.6 min (3 blocks per stimulation type x 32 sec)  Behavioural: 39.37–50.35 min | fMRI: 1 (9 stimulation blocks: 3 blocks x 3 stimulation types)  Behavioural: 2 (1 tTIS, 1 sham) |
| Wessel et al. (2023) | fMRI: tbfMRI 30 min (6 blocks per stimulation type x 30 sec x 10 repetitions), rsfMRI 8 min  Behavioural: 10.5 min (7 blocks x 90 sec) | fMRI: 4 (2 rsfMRI, 2 tbfMRI)  Behavioural: 2 (1 tTIS, 1 tACS) |
| Beanato et al. (2024) | 18 min (2 blocks per stimulation type x 9 min) | 1 (2 blocks iTBS tTIS, 2 blocks cTBS tTIS, 2 blocks sham) |
| Demchenko et al. (2024) | 30 min | 10 (1x/day for 2 weeks) |
| Liu et al. (2024) | 10 min (on-line effect); 20 min (off-line effect) | 1-2 (1 patient completed tTIS and tACS, 2 patients completed tTIS only) |
| Modak et al. (2024) | 16 min (2 blocks per stimulation type x 8 min) | 1 (2 tTIS blocks, 2 sham blocks) |
| Thiele et al. (2024) | 20 min (10 min resting block + 10 min mental rotation task) | 1 (1 resting-state block, 1 mental rotation task block) |
| Vassiliadis et al. (2024a)^a^ | 10 min (2 blocks per stimulation type x 5 min) | 1 (2 blocks tTIS 20 Hz, 2 blocks tTIS 80 Hz, 2 blocks sham) |
| Vassiliadis et al. (2024b)^b^ | 7–40 min | 1–4 (257 sessions total) |
| Wang et al. (2024) | 30 min | 1 |
| Yang et al. (2024a)^c^ | 20 min | 2 (1 tTIS, 1 sham) |
| Yang et al. (2024b)^d^ | 20 min | 1 |
| Zheng et al. (2024) | 20 min | 10 (2x day for 5 days) |
| Zhu et al. (2024) | 20 min | 2 (1 tTIS, 1 tDCS) |
| **Abbreviations:** **cTBS** = Continuous theta burst stimulation; **fMRI** = Functional magnetic resonance imaging; **iTBS** = Intermittent theta burst stimulation; **rsfMRI** = Resting-state functional magnetic resonance imaging; **tACS** = Transcranial alternating current stimulation; **tbfMRI** = Task-based functional magnetic resonance imaging; **tDCS** = Transcranial direct current stimulation; **tTIS** = Transcranial temporal interference stimulation; **x** = Times (e.g., 3 blocks × 10 min).  ^a^ Vassiliadis et al. (2024). *J Neural Eng*.  ^b^ Vassiliadis et al. (2024). *Nat Hum Behav*.  ^c^ Yang et al. (2024). *Mov Disord*.  ^d^ Yang et al. (2024). *Brain Stimul*. | | |

###### eTable S8. Summary of Stimulation Parameters in Ongoing tTIS Clinical Trials

| **Study** | **Brain target** | **tTIS montage** | **Envelope frequency, *Δf*** | **Carrier frequencies,**  ***f_1_ \| f_1_ + Δf*** | **Zero-to-peak amplitude,**  ***I_1_* \| *I_2_*** | **Ramp time** | **Number of sessions per participant** | **tTIS device manufacturer** |
| --- | --- | --- | --- | --- | --- | --- | --- | --- |
| NCT03747601  (2018) | V1 (calcarine cortex) | ns | ns | ns | ns | ns | 2-4 | ns |
| NCT06716866  (2019) | ns | ns | ns | ns | ns | ns | ns | ns |
| NCT04099056  (2019) | ns | ns | ns | ns | ns | ns | ns | ns |
| NCT04432064  (2020) | ns | ns | ns | ns | ns | 60 min | 1 or 5 | ns |
| NCT04761471  (2021) | ns | ns | ns | ns | ns | <30 min | ns | ns |
| NCT05777876  (2023) | Identified via data-driven methods | ns | ~130 Hz (20 Hz for HC) | ns | ns | 30 min | 10 | ns |
| NCT05805215  (2023) | Hippocampus, precuneus | ns | <100 Hz | ns | ns | ns | ns | ns |
| ChiCTR2300077258 (2023) | ns | ns | 130 Hz | 1300 Hz \| 1430 Hz | 2.5 mA \| 2 mA | 20 min | 2 | Soterix Medical, NJ, USA |
| DRKS00030841  (2023) | Putamen, NAc | ns | ns | ns | ns | ns | ns | ns |
| JPRN-UMIN000050056 (2023) | Insula, S1 | ns | ns | ns | ns | ns | ns | ns |
| NCT06663969  (2024) | Deep brain nuclei | ns | ns | ns | ns | ns | ns | ns |
| NCT06708143  (2024) | Deep brain nuclei | ns | ns | ns | ns | ns | ns | ns |
| ChiCTR2400090858 (2024) | ns | ns | 10 Hz | 2000 Hz \| 2010 Hz | 2 mA \| 2 mA | 20 min | 40 | ns |
| ChiCTR2400090777 (2024) | vmPFC | Fp1-F5 & Fp2-AF8 | 10 Hz; 5 Hz | 2000 Hz \| 2010 Hz; 2000 Hz \| 2005 Hz | 0.85 mA \| 1.15 mA | 20 min | 3 | Soterix Medical, NJ, USA |
| ChiCTR2400085831 (2024) | ns | ns | ns | ns | ns | 20 min | 10 | ns |
| ChiCTR2400090107 (2024) | GPi, putamen | ns | 10 Hz | 2000 Hz \| 2010 Hz | 2 mA \| 2 mA | 20 min | 20 | ns |
| ChiCTR2400090199 (2024) | Left dlPFC, left hippocampus | ns | ns | ns | 2 mA \| 2 mA | 30 min | 20 | ns |
| NCT06452849  (2024) | dlPFC, amygdala | ns | 10 Hz (dlPFC); 100 Hz (amygdala) | ns | ns | 20 min | 5 | NeuroDome Medical Technology Co., Xi'an, Shaanxi, China |
| NCT06461260  (2024) | Amygdala | ns | 100 Hz | ns | ns | 20 min | 5 | NeuroDome Medical Technology Co., Xi'an, Shaanxi, China  ns |
| NCT06467422  (2024) | dACC | ns | 6 Hz | 2000 Hz \| 2006 Hz | 2 mA \| 2 mA | 20 min | 1 |  |
| NCT06477276  (2024) | Right amygdala | ns | ns | ns | ns | ns | 5 | NeuroDome Medical Technology Co., Xi'an, Shaanxi, China |
| NCT06516991  (2024) | NAc | ns | ns | ns | 3.64 mA \| 4.36 mA | 30 min | 10 | ns |
| NCT06547086  (2024) | ns | ns | ns | ns | ns | ns | ns | ns |
| NCT06601686  (2024) | PMC | ns | ns | ns | ns | ns | ns | ns |
| NCT06601699  (2024) | PMC | ns | ns | ns | ns | 5 min | Up to 24 (Up to 8x daily, 3 days separated by one week each) | ns |
| NCT06607432  (2024) | Pulvinar nucleus | ns | ns | ns | ns | ns | ns | ns |
| NCT06267521  (2024) | ns | ns | ns | ns | ns | 10 hours/night | 4 or 8 | ns |
| ChiCTR2400081207 (2024) | ns | ns | ns | ns | ns | 25 min | 10 | ns |
| **Abbreviations:** dACC = Dorsal anterior cingulate cortex; dlPFC = Dorsolateral prefrontal cortex; GPi = Globus pallidus internus; HC = Healthy control; NAc = Nucleus accumbens; ns = Not specified; PMC = Posteromedial cortex; S1 = Primary somatosensory cortex; V1 = Primary visual cortex; vmPFC = Ventromedial prefrontal cortex. | | | | | | | | |

###### eTable S9. Clinical Outcomes from Controlled tTIS Trials in Parkinson’s Disease and Essential Tremor

| **Study** | **Target** | **Sample size** | **tTIS vs. Control** | **Evidence for Efficacy** |
| --- | --- | --- | --- | --- |
| Yang et al. (2024) | Right GPi | 12 | Superior | Preliminary |
| Liu et al. (2024) | Bilateral SN | 1 | Superior*** | Inconclusive |

**Abbreviations:** GPi = Globus pallidus internus; SN = Substantia nigra.

Evidence for efficacy was categorized based on the framework adapted from Heim et al. (2023). Preliminary: One available study demonstrating superior efficacy. Inconclusive: No available studies demonstrating efficacy of tTIS, or data are insufficient to make a determination.

In Liu et al. (2024), 2 of the 3 participants (Essential Tremor: *n*=1, Parkinson’s disease: *n*=1) were excluded, as they did not complete the control condition due to fatigue or scheduling constraints. These participants exhibited a reduction in continuity of resting tremor for the active tTIS condition relative to baseline. For other assessment items, the overall direction of change could not be determined.

Yang et al. (2024). *Mov Disord*.

###### eTable S10. Effect Sizes for Clinical Outcomes in an Open-Label tTIS Trial in Parkinson’s Disease

|  | **Pre-Post (Immediate)** | | **Pre-Post (30-min)** | | **Pre-Post (60-min)** | |
| --- | --- | --- | --- | --- | --- | --- |
| **Outcome** | **Hedge’s *g***  **(95% CI)** | **MD**  **(95% CI)** | **Hedge’s *g***  **(95% CI)** | **MD**  **(95% CI)** | **Hedge’s *g***  **(95% CI)** | **MD**  **(95% CI)** |
| Total | -0.92  (-1.49, -0.35) | -11.12  (-17.50, -4.75) | -0.88  (-1.30, -0.46) | -11.88  (-17.21, -6.54) | -0.73  (-1.17, -0.30) | -10.12  (-16.03, -4.22) |
| Rigidity | -0.88  (-1.35, -0.41) | -2.88  (-4.32, -1.43) | -0.66  (-1.18, -0.13) | -2.12  (-3.82, -0.43) | -0.71  (-1.17, -0.26) | -2.38  (-3.85, -0.90) |
| Bradykinesia | -0.72  (-1.38, -0.05) | -4.75  (-9.08, -0.42) | -0.75  (-1.25, -0.24) | -5.00  (-8.28, -1.72) | -0.76  (-1.56, 0.04) | -4.62  (-9.37, 0.12) |
| Axial | -0.28  (-0.57, 0.00) | -1.12  (-2.34, 0.09) | -0.34  (-0.52, -0.17) | -1.62  (-2.51, -0.74) | -0.39  (-0.63, -0.14) | -1.62  (-2.71, -0.54) |
| Tremor | -0.35  (-0.88, 0.18) | -2.38  (-6.27, 1.52) | -0.45  (-0.93, 0.04) | -3.12  (-6.71, 0.46) | -0.17  (-0.48, 0.14) | -1.50  (-4.56, 1.56) |
| **Side** | | | | | | |
| Ipsilateral | -1.06  (-2.09, -0.03) | -3.75  (-6.96, -0.54) | -1.04  (-1.62, -0.45) | -4.12  (-6.19, -2.06) | -0.54  (-0.95, -0.12) | -2.75  (-4.93, -0.57) |
| Contralateral | -0.44  (-0.66, -0.22) | -4.88  (-7.42, -2.33) | -0.42  (-0.70, -0.14) | -4.00  (-6.83, -1.17) | -0.41  (-0.83, 0.01) | -3.38  (-7.06, 0.31) |

**Abbreviations:** CI = Confidence Interval; MD = Mean Difference; MDS-UPDRS-III = Movement Disorder Society Unified Parkinson's Disease Rating Scale Part III**.**

Hedge’s *g* was derived using the standard deviation of differences and Hedge’s correction factor. MDs and CIs were computed using the *t*-distribution with *n*–1 degrees of freedom.

Yang et al. (2024). *Brain Stimul*.

###### eTable S11. Behavioural Outcomes from Controlled tTIS Trials in Healthy Participants

| **Behavioural Domains** | **Superior to control (*n*, %)** | **No difference (*n*, %)** | **Evidence for Efficacy** |
| --- | --- | --- | --- |
| All studies, *n* = 13 | 6 (46%) | 7 (54%) | Indicative |
| Cognitive Function, *n* = 3 | 0 (0%) | 3 (100%) | Inconclusive |
| Memory, *n* = 2 | 2 (100%) | 0 (0%) | Indicative |
| Visual, *n* = 2 | 0 (0%) | 2 (100%) | Inconclusive |
| Motor, *n* = 6 | 4 (67%) | 2 (33%) | Indicative |

Evidence for efficacy was categorized based on the framework adapted from Heim et al. (2023). Indicative: ≥2 available RCTs with ≥50% demonstrating superior efficacy. Inconclusive: ≥1 available RCTs with >50% not demonstrating superior efficacy.

*n* denotes the number of studies.

###### eTable S12. Effect Sizes for Human tTIS Studies Reporting Neuroimaging and Neurophysiology Outcomes

| **Study** | **Modality** | **Target/Region** | **MRI/EEG Effect Size** | **Magnitude** |
| --- | --- | --- | --- | --- |
| Beanato et al. (2024) | fMRI (VR navigation) | Right hippocampal–entorhinal complex | Entorhinal GCLR: iTBS < Control (*p*<0.001), cTBS < Control (*p*=0.019), iTBS < cTBS (*p*=0.030);  Control GCLR: *r*=0.48 (*p*=0.009), iTBS *r*=0.42 (ns), cTBS *r*=0.049 (ns). Hippocampal BOLD Cue+Retrieval > EC (*t*(319)=-5.155, *p*<0.001, *d*=-0.77); Hipp BOLD-behaviour correlation *r*=-0.55 (*p*=0.01) | Medium-Large (GCLR, BOLD–behavior) |
| Iszak et al. (2023) | EEG, phosphene, EMG | Visual cortex, motor cortex | EEG: No alpha ↑ with tTIS (*Δ*≈-0.94 dB, ns); Friedman *χ²(*3)=33.56, *p*<0.001 driven by tACS (*Δ*≈+0.67 dB). Phosphenes: induced by tACS (~100 µA), not by tTIS (≤4000 µA). Peripheral EMG: reliable muscle twitches at ~1.2–2.3 mA; thresholds correlated with body weight (*ρ*=0.56, *p*=0.01) | Null-Small (EEG, phosphenes); Large (peripheral) |
| Modak et al. (2024) | fMRI (resting state) | Left caudate (target), OFC, parahippocampal gyrus | tTIS-Sham contrast: Fusiform/parahippocampal cluster *t*=6.31, *p*=0.011 (FWE corr); Mid-OFC cluster *t*=5.10, *p*=0.092 (trend), SVC *p*=0.021; Left caudate ROI ns (*p*=0.295). Logistic fit suggested dose-response relation | Medium-Large (parahippocampal, OFC) |
| Thiele et al. (2024) | EEG (mental rotation) | Parietal cortex (ERD, alpha power) | ERD effect: *F*=4.80, *p*=0.013, *η²_p_*=0.176 | Medium |
| Vassiliadis et al. (2024) | fMRI + motor learning | Striatum (putamen, caudate) | Striatal BOLD: Reinforcement ON > OFF (*p*<0.01), no main effect of tTIS type. Correlation: (tTIS_80Hz_–tTIS_20Hz_) modulation correlated with RL ability (clusters in L putamen, bilateral caudate; voxel *p*=0.001, cluster-FDR *p*=0.05). Effective connectivity: gPPI showed ↑ striatum→frontal cortex under tTIS_80Hz_+Reinf (*p*=0.001 vs ReinfOFF; *p*<0.001 vs sham) | Medium-Large (connectivity, correlations) |
| Violante et al. (2023) | fMRI (working memory) | Right hippocampus, fronto-parietal network | Encoding BOLD: left hippocampus *t*(19)=3.70, *p*=0.003; right hippocampus *t*(19)=3.92, *p*=0.003 (FDR corr). tTIS 1:3 reduced left hippocampal BOLD vs sham (*F*(2,95)=3.2, *p*=0.0443; post-hoc sham vs tTIS 1:3 *p*=0.006). Voxelwise: anterior ~30% and posterior ~25% hippocampal voxels ↓ (TFCE FWE *p*<0.05). Connectivity: hippocampal–AT network ↓ (*F*(4,1576)=2.54, p=0.038) | Medium (cluster-level, connectivity) |
| Wessel et al. (2023) | fMRI + motor task | Putamen, SMA, thalamus, cerebellum | ROI: Putamen > caudate (*F*(1,276)=260.01, *η²_p_* =0.49); Region×stimulation *F*(1,276)=4.48, *p*=0.035, *η²_p_* =0.02. Putamen activation ↑ with tTIS vs HF-tACS (*t*(276)=-2.55, *p*=0.01, *d*=-0.41). Subregions: *F*(1,299)=13.47, *p*=0.0003, *η²_p_* =0.04; posterior > anterior *F*(1,299)=37.03, *p*=3.6e-9, *η²_p_* =0.11. Connectivity (PPI putamen→motor): *F*(2,450)=30.70, *p*=3.2e-13, *η²_p_* =0.12; stim×cluster *F*(2,450)=3.26, *p*=0.04, *η²_p_* =0.01. Voxelwise: ↑ BOLD in striatum, thalamus, SMA, cerebellum (FDR corr) | Medium (ROI, connectivity, voxelwise) |
| Zhu et al. (2022) | fMRI (resting state) | Left M1 → premotor, SMA, SFG, MFG, SPL | Cluster 1: 1078 vox, *Z*=4.40, *p*<0.05 FWE corr; Cluster 2: 218 vox, *Z*=4.13, *p*<0.05 FWE corr | Medium-Large (cluster-level) |
| Zhu et al. (2024) | fMRI (dynamic FC) | Left M1 (FDI hotspot, pre/postcentral gyri) | dFC CV reduced: *t*=3.57, *p*<0.001; Mean dFC increased: *t*=4.16, *p*=0.018; Within-tTIS baseline vs online: *t*=6.58 (all significant) | Medium-Large (t-statistics) |
| von Conta et al. (2022) | MEG (alpha power) | Parieto-occipital cortex | Alpha power ↑ pre→post in all groups (tTIS *V*=77, *p*<0.01; tACS *V*=111, *p*<0.01; Control *V*=96, *p*<0.01). No between-condition differences: tTIS vs control *V*=226, *p*=0.17, BF_10_=0.52; tACS vs control *V*=304, *p*=0.66, BF_10_=0.16; tTIS vs tACS *V*=204, *p*=0.18, BF_10_=0.45 | Null-Small (no condition differences) |

**Abbreviations:** BOLD = blood oxygen level–dependent signal; dFC = dynamic functional connectivity; ERD = event-related desynchronization; ERS = event-related synchronization; FDR = false discovery rate; FWE = family-wise error; gPPI = generalized psychophysiological interaction; GCLR = grid cell-like representation; HF = high-frequency control; M1 = primary motor cortex; MEG = magnetoencephalography; ns = not significant; OFC = orbitofrontal cortex; ROI = region of interest; SMA = supplementary motor area; SVC = small volume correction; tACS = transcranial alternating current stimulation; tDCS = transcranial direct current stimulation; tTIS = transcranial temporal interference stimulation.

Piao et al. (2022) and Wang et al. (2024) collected EEG data for safety monitoring only, and no effect sizes were reported. Demchenko et al. (2024) is a protocol paper with no results reported.

### PRISMA 2020 Checklist for Abstracts

| **Section and Topic** | **Item #** | **Checklist item** | **Reported (Yes/No)** |
| --- | --- | --- | --- |
| **TITLE** | | |  |
| Title | 1 | Identify the report as a systematic review. | Yes |
| **BACKGROUND** | | |  |
| Objectives | 2 | Provide an explicit statement of the main objective(s) or question(s) the review addresses. | Yes |
| **METHODS** | | |  |
| Eligibility criteria | 3 | Specify the inclusion and exclusion criteria for the review. | Yes |
| Information sources | 4 | Specify the information sources (e.g. databases, registers) used to identify studies and the date when each was last searched. | Yes |
| Risk of bias | 5 | Specify the methods used to assess risk of bias in the included studies. | Yes |
| Synthesis of results | 6 | Specify the methods used to present and synthesise results. | No (reported in the body) |
| **RESULTS** | | |  |
| Included studies | 7 | Give the total number of included studies and participants and summarise relevant characteristics of studies. | Yes |
| Synthesis of results | 8 | Present results for main outcomes, preferably indicating the number of included studies and participants for each. If meta-analysis was done, report the summary estimate and confidence/credible interval. If comparing groups, indicate the direction of the effect (i.e. which group is favoured). | Yes |
| **DISCUSSION** | | |  |
| Limitations of evidence | 9 | Provide a brief summary of the limitations of the evidence included in the review (e.g. study risk of bias, inconsistency and imprecision). | No (reported in the body) |
| Interpretation | 10 | Provide a general interpretation of the results and important implications. | Yes |
| **OTHER** | | |  |
| Funding | 11 | Specify the primary source of funding for the review. | N/A |
| Registration | 12 | Provide the register name and registration number. | Yes |

*From:* Page MJ, McKenzie JE, Bossuyt PM, Boutron I, Hoffmann TC, Mulrow CD, et al. The PRISMA 2020 statement: an updated guideline for reporting systematic reviews. BMJ 2021;372:n71. doi: 10.1136/bmj.n71. This work is licensed under CC BY 4.0. To view a copy of this license, visit <https://creativecommons.org/licenses/by/4.0/>

### PRISMA 2020 Checklist

| **Section and Topic** | **Item #** | **Checklist item** | **Location where item is reported** |
| --- | --- | --- | --- |
| **TITLE** | | |  |
| Title | 1 | Identify the report as a systematic review. | Title |
| **ABSTRACT** | | |  |
| Abstract | 2 | See the PRISMA 2020 for Abstracts checklist. | Abstract |
| **INTRODUCTION** | | |  |
| Rationale | 3 | Describe the rationale for the review in the context of existing knowledge. | Introduction, paragraphs 1-3 |
| Objectives | 4 | Provide an explicit statement of the objective(s) or question(s) the review addresses. | Introduction, paragraph 3 |
| **METHODS** | | |  |
| Eligibility criteria | 5 | Specify the inclusion and exclusion criteria for the review and how studies were grouped for the syntheses. | Methods (*Study Selection*), Supplement (*Eligibility Criteria*, *eResults* p. 12) |
| Information sources | 6 | Specify all databases, registers, websites, organisations, reference lists and other sources searched or consulted to identify studies. Specify the date when each source was last searched or consulted. | Methods (*Search Strategy*) |
| Search strategy | 7 | Present the full search strategies for all databases, registers and websites, including any filters and limits used. | Supplement (*Search Strategy,* pp. 2-4) |
| Selection process | 8 | Specify the methods used to decide whether a study met the inclusion criteria of the review, including how many reviewers screened each record and each report retrieved, whether they worked independently, and if applicable, details of automation tools used in the process. | Methods (*Study Selection*) |
| Data collection process | 9 | Specify the methods used to collect data from reports, including how many reviewers collected data from each report, whether they worked independently, any processes for obtaining or confirming data from study investigators, and if applicable, details of automation tools used in the process. | Methods (*Data Extraction and Appraisal of Methodological Quality*), Supplement (*List of Extracted Variables*) |
| Data items | 10a | List and define all outcomes for which data were sought. Specify whether all results that were compatible with each outcome domain in each study were sought (e.g. for all measures, time points, analyses), and if not, the methods used to decide which results to collect. | Methods (*Data Extraction and Appraisal of Methodological Quality*), Supplement (*List of Extracted Variables*) |
|  | 10b | List and define all other variables for which data were sought (e.g. participant and intervention characteristics, funding sources). Describe any assumptions made about any missing or unclear information. | Methods (*Data Extraction and Appraisal of Methodological Quality*), Supplement (*List of Extracted Variables*) |
| Study risk of bias assessment | 11 | Specify the methods used to assess risk of bias in the included studies, including details of the tool(s) used, how many reviewers assessed each study and whether they worked independently, and if applicable, details of automation tools used in the process. | Methods (*Data Extraction and Appraisal of Methodological Quality*) |
| Effect measures | 12 | Specify for each outcome the effect measure(s) (e.g. risk ratio, mean difference) used in the synthesis or presentation of results. | Methods (*Statistical Analysis*) |
| Synthesis methods | 13a | Describe the processes used to decide which studies were eligible for each synthesis (e.g. tabulating the study intervention characteristics and comparing against the planned groups for each synthesis (item #5)). | Methods (*Statistical Analysis*), Supplement (*eResults* p.12) |
|  | 13b | Describe any methods required to prepare the data for presentation or synthesis, such as handling of missing summary statistics, or data conversions. | Methods (*Statistical Analysis*), Supplement (eTables S9-12) |
|  | 13c | Describe any methods used to tabulate or visually display results of individual studies and syntheses. | Methods (*Statistical Analysis*) |
|  | 13d | Describe any methods used to synthesize results and provide a rationale for the choice(s). If meta-analysis was performed, describe the model(s), method(s) to identify the presence and extent of statistical heterogeneity, and software package(s) used. | Methods (*Statistical Analysis*), Supplement (*eResults* p. 12) |
|  | 13e | Describe any methods used to explore possible causes of heterogeneity among study results (e.g. subgroup analysis, meta-regression). | N/A |
|  | 13f | Describe any sensitivity analyses conducted to assess robustness of the synthesized results. | N/A |
| Reporting bias assessment | 14 | Describe any methods used to assess risk of bias due to missing results in a synthesis (arising from reporting biases). | Methods (*Data Extraction and Appraisal of Methodological Quality*) |
| Certainty assessment | 15 | Describe any methods used to assess certainty (or confidence) in the body of evidence for an outcome. | Methods (*Data Extraction and Appraisal of Methodological Quality*) |
| **RESULTS** | | |  |
| Study selection | 16a | Describe the results of the search and selection process, from the number of records identified in the search to the number of studies included in the review, ideally using a flow diagram. | Results (*Study Selection*), Figure 1 |
|  | 16b | Cite studies that might appear to meet the inclusion criteria, but which were excluded, and explain why they were excluded. | N/A |
| Study characteristics | 17 | Cite each included study and present its characteristics. | References, Table 1, Supplement (eTable S4) |
| Risk of bias in studies | 18 | Present assessments of risk of bias for each included study. | Results (*Methodological Quality of Studies*), Supplement (eTable S1, eTable S2) |
| Results of individual studies | 19 | For all outcomes, present, for each study: (a) summary statistics for each group (where appropriate) and (b) an effect estimate and its precision (e.g. confidence/credible interval), ideally using structured tables or plots. | Table 1-2, Supplement (eTables S4-12) |
| Results of syntheses | 20a | For each synthesis, briefly summarise the characteristics and risk of bias among contributing studies. | Results (*Research Design and Participants*, *Methodological Quality of Studies*), Supplement (*eResults* p. 11) |
|  | 20b | Present results of all statistical syntheses conducted. If meta-analysis was done, present for each the summary estimate and its precision (e.g. confidence/credible interval) and measures of statistical heterogeneity. If comparing groups, describe the direction of the effect. | Results (*Brain Targets and Stimulation Parameters, Safety and Tolerability Outcomes, Clinical Outcomes, Behavioural Outcomes, Neuroimaging Outcomes, Neurophysiological Outcomes*), Supplement (*eResults* pp. 11-15, eTables S9-12) |
|  | 20c | Present results of all investigations of possible causes of heterogeneity among study results. | N/A |
|  | 20d | Present results of all sensitivity analyses conducted to assess the robustness of the synthesized results. | N/A |
| Reporting biases | 21 | Present assessments of risk of bias due to missing results (arising from reporting biases) for each synthesis assessed. | Results (*Methodological Quality of Studies*), Supplement (eTable S1, eTable S2) |
| Certainty of evidence | 22 | Present assessments of certainty (or confidence) in the body of evidence for each outcome assessed. | Supplement (eTable S3) |
| **DISCUSSION** | | |  |
| Discussion | 23a | Provide a general interpretation of the results in the context of other evidence. | Discussion |
|  | 23b | Discuss any limitations of the evidence included in the review. | Discussion (*Strengths and Limitations*) |
|  | 23c | Discuss any limitations of the review processes used. | Discussion (*Strengths and Limitations*) |
|  | 23d | Discuss implications of the results for practice, policy, and future research. | Conclusions |
| **OTHER INFORMATION** | | |  |
| Registration and protocol | 24a | Provide registration information for the review, including register name and registration number, or state that the review was not registered. | Abstract, Methods (*Search Strategy*), PROSPERO |
|  | 24b | Indicate where the review protocol can be accessed, or state that a protocol was not prepared. | PROSPERO |
|  | 24c | Describe and explain any amendments to information provided at registration or in the protocol. | PROSPERO |
| Support | 25 | Describe sources of financial or non-financial support for the review, and the role of the funders or sponsors in the review. | Funding |
| Competing interests | 26 | Declare any competing interests of review authors. | Declaration of competing interests |
| Availability of data, code and other materials | 27 | Report which of the following are publicly available and where they can be found: template data collection forms; data extracted from included studies; data used for all analyses; analytic code; any other materials used in the review. | Data Availability |

*From:* Page MJ, McKenzie JE, Bossuyt PM, Boutron I, Hoffmann TC, Mulrow CD, et al. The PRISMA 2020 statement: an updated guideline for reporting systematic reviews. BMJ 2021;372:n71. doi: 10.1136/bmj.n71. This work is licensed under CC BY 4.0. To view a copy of this license, visit <https://creativecommons.org/licenses/by/4.0/>
